# Modeling transmission dynamics and effectiveness of worker screening programs for SARS-CoV-2 in pork processing plants

**DOI:** 10.1101/2021.03.02.21249552

**Authors:** Kimberly VanderWaal, Lora Black, Judy Hodge, Addisalem Bedada, Scott Dee

## Abstract

Pork processing plants were apparent hotspots for SARS-CoV2 in the spring of 2020. As a result, the swine industry was confronted with a major occupational health, financial, and animal welfare crisis. The objective of this work was to describe the epidemiological situation within processing plants, develop mathematical models to simulate transmission in these plants, and test the effectiveness of routine PCR screening at minimizing SARS-CoV2 circulation. Cumulative incidence of clinical (PCR-confirmed) disease plateaued at ∼2.5% to 25% across the three plants studied here. For larger outbreaks, antibody prevalence was approximately 30% to 40%. Secondly, we developed a mathematical model that accounts for asymptomatic, pre-symptomatic, and background “community” transmission. By calibrating this model to observed epidemiological data, we estimated the initial reproduction number (*R*) of the virus. Across plants, *R* generally ranged between 2 and 4 during the initial phase, but subsequently declined to ∼1 after two to three weeks, most likely as a result of implementation/compliance with biosecurity measures in combination with population immunity. Using the calibrated model to simulate a range of possible scenarios, we show that the effectiveness of routine PCR-screening at minimizing disease spread was far more influenced by testing frequency than by delays in results, *R*, or background community transmission rates. Testing every three days generally averted about 25% to 40% of clinical cases across a range of assumptions, while testing every 14 days typically averted 7 to 13% of clinical cases. However, the absolute number of additional clinical cases expected and averted was influenced by whether there was residual immunity from a previous peak (i.e., routine testing is implemented after the workforce had experienced an initial outbreak). In contrast, when using PCR-screening to prevent outbreaks or in the early stages of an outbreak, even frequent testing may not prevent a large outbreak within the workforce. This research helps to identify protocols that minimize risk to occupational safety and health and support continuity of business for U.S. processing plants. While the model was calibrated to meat processing plants, the structure of the model and insights about testing are generalizable to other settings where large number of people work in close proximity.

## Introduction

Almost 9000 U.S. meat, poultry, and agricultural workers tested positive for SARS-CoV2 from March through May 2020 (1), and at the time, the numerous outbreaks that occurred at meat processing plants were amongst the largest and fastest growing workplace outbreaks in the country (2). In addition, disease incidence appears to be elevated in communities in proximity to processing plants, suggesting that the presence of plants may amplify transmission in the surrounding population (3). In some cases, >25% of workers tested positive (4), leading to plant closures or reduced operations affecting 25% of U.S. meat processing capacity during April and May (2). As a result, the swine industry was confronted with a major occupational health, financial, and animal welfare crisis; at the time, it was feared that up to 700,000 pigs would need to be culled per week due to stalled and backlogged production chains (5), and U.S. meat production fell by 20% by the end of April (6). Plants largely returned to normal capacity by June (7). However, similar to other high-density places of work and study, worker health and safety remains an ongoing concern.

Meat processing plants may be particularly susceptible to spread of respiratory diseases due to both environmental and human conditions (8), such as prolonged contact amongst employees working in high density settings. This is often further compounded by shared transportation and accommodation, as well as socializing outside of work (1). Between March 1 and May 31 2020, 8,978 cases and 55 deaths were reported across 742 meat, poultry, and crop production workplaces in 30 states (1). Testing strategies varied by workplace, and not all workplaces performed mass testing (1). In addition, community-based testing was quite limited during this period, so the true number of cases in such workplaces relative to background rates of community transmission is largely uncertain. While continued operation of processing facilities is essential for food supply chains, analysis of county-level data reveals a worrisome trend in which counties with processing plants reported >200,000 excess cases and > 4,000 excess deaths compared to nearby counties, with the majority of spread likely attributed to community transmission, but amplified by the presence of a plant (3). Similar situations have also been reported in Germany, Portugal, and the United Kingdom (9). Data related to this issue have thus far been reported in an aggregated manner (1, 3, 10), and there are limited data and epidemiological analysis of how outbreaks unfolded at individual plants (but see (4).

To better understand how outbreaks can be controlled or prevented, screening/testing protocols have been proposed to identify infected and asymptomatic individuals, often with an emphasis on health care environments (11, 12), high-risk cohorts (13), and large populations (14). For example, weekly screening of high-risk populations (such as healthcare workers) can reduce their contribution to disease spread by ∼25% relative to a strategy based upon isolation of symptomatic cases alone (13). However, the population-level effectiveness of different screening/testing protocols, such as screening asymptomatic workers on a routine basis, has not been rigorously tested for non-healthcare workplaces.

Working in close partnership with industry stakeholders, the objective of this work was to first characterize the epidemic curves across three pork processing plants and summarize epidemiological data related to PCR and antibody testing at individual plants. We then develop a mathematical model to simulate the spread of SARS-CoV2 within these plants, accounting for pre-symptomatic, asymptomatic, and community transmission, and calibrate the model to observed epidemiological data. We used this model to estimate each outbreaks’ reproduction number (*R*) during the early phases of workplace transmission and calculated time-dependent reproduction numbers (*R*-TD) that allow the effective R to vary through time to measure the influence of interventions (15). Finally, we used the model to evaluate a range of PCR-screening scenarios to test their effectiveness at minimizing SARS-CoV2 circulation.

## Methods

### Study populations and descriptive epidemiology

Data were available from three plants located in different states. These plants ranged in size from 750 workers to 2400 workers. Plant attributes are summarized in Table 1. Plants participated in this study on condition of confidentiality, therefore company names and plant locations are not identified in this paper. Each plant had two types of data available: daily incidence of new self-reported SARS-CoV2 cases and company-initiated testing data. Each plant became involved in this project during the early stages of COVID-19 as part of a cooperative needs assessment for the swine industry, which was a newly formed interdisciplinary group of swine veterinarians, epidemiologists, medical doctors, and diagnosticians that contributed to the development of the model and interpretation of results. The driver of this novel collaboration was an intrinsic understanding of the impact that this human health problem would have on animal welfare and food production. As such, the plants included in this project were not randomly selected and were aggressive and progressive in implementing control measures for COVID-19 early in the pandemic.

**Table 1.** Plant attributes and results of company-initiated testing, including timing, sample size, and the percent positive to PCR and antibody testing. 95% confidence intervals are shown in parenthesis.

| Plant | Workforce size | Number shifts | Plant age | Day* | No. tested | IgM | IgG |
| --- | --- | --- | --- | --- | --- | --- | --- |
| A | 1128 | 1 | <5 years | 9 | 298 | 9.4%<br>(6.1-12.7%) | 6.4%<br>(3.6-9.2%) |
| B | 2400 | 2 | >50 years | 46 | 472 | 19.0%<br>(15.4-22.5%) | 35.8%<br>(31.5-40.1%) |
| C | 750 | 1 | <5 years | 25 | 392 |  | 16.6%<br>(12.9-20.3%) |
|  |  |  |  | 81 | 218 |  | 29.8%<br>(23.7-35.9%) |

\* Day testing concluded (counted from earliest date with 5 cumulative cases)

### Daily incidence of clinical cases

Daily incidence of self-reported cases was summarized as the number of new workers per day who reported a confirmed SARS-CoV2 positive PCR test administered by their private health care provider (Plants A – B) or through onsite company-sponsored testing (Plant C). Given that testing by health care providers was almost exclusively limited to symptomatic cases during this time period, self-reported cases were assumed to represent the incidence of clinical disease and not pre-symptomatic or asymptomatic infections (see supplementary methods S1 for further detail). From these datasets, we plotted the epidemic curve for each plant based on daily clinical cases as well as cumulative clinical cases (plotted as a proportion to facilitate comparisons between plants). Given that the very early reporting of clinical cases may be subject to error, epidemic curves were plotted relative to days elapsed since the first five clinical cases.

### Company-initiated testing

All plants underwent company-initiated testing of workers, though testing strategy varied between plants (Table 1). Plants A and B tested a randomly selected subset of the workforce with an antibody test (IgG/IgM Rapid Test Cassette, Healgen Scientific, Houston, TX). The manufacturer reports the test’s positive percent agreement as 86.7% and 96.7% for IgM and IgG respectively. Negative percent agreement was 99.0% and 98.0% for IgM and IgG, respectively. Plant C also employed antibody tests (LIAISON® SARS-CoV-2 S1/S2 IgG, DiaSorin Inc, Stillwater, Minnesota,). Reported test performance for IgG was 97.6% positive percent agreement and 99.3% negative percent agreement at >15 days post-infection. Testing was conducted on a single date at Plant A, spread over a series of weeks in Plant B (Supplementary Table S2), and on two distinct dates separated by ∼2 months in Plant C. For all three plants, selection of tested workers was pseudo-random on a voluntary basis, and workers were asked about their infection history (presence of clinical signs, symptom onset, PCR testing results) at the time of testing. Plant C provided on-site PCR-testing on a rolling basis.

### Model framework

We developed deterministic and stochastic compartmental models to simulate the transmission of SARS-CoV2 within plants, assuming homogenous mixing, which was deemed a reasonable assumption based on discussions with plant representatives and given the small scale of the model. We used the deterministic version of the model for the purposes of model-calibration to minimize computational time. However, the stochastic version of the model allowed us to better quantify variability in model outcomes, as well as introduce events into the model that only occurred at certain intervals (for example, worker screening/testing programs, red lines in Figure 1).

**Figure 1.**
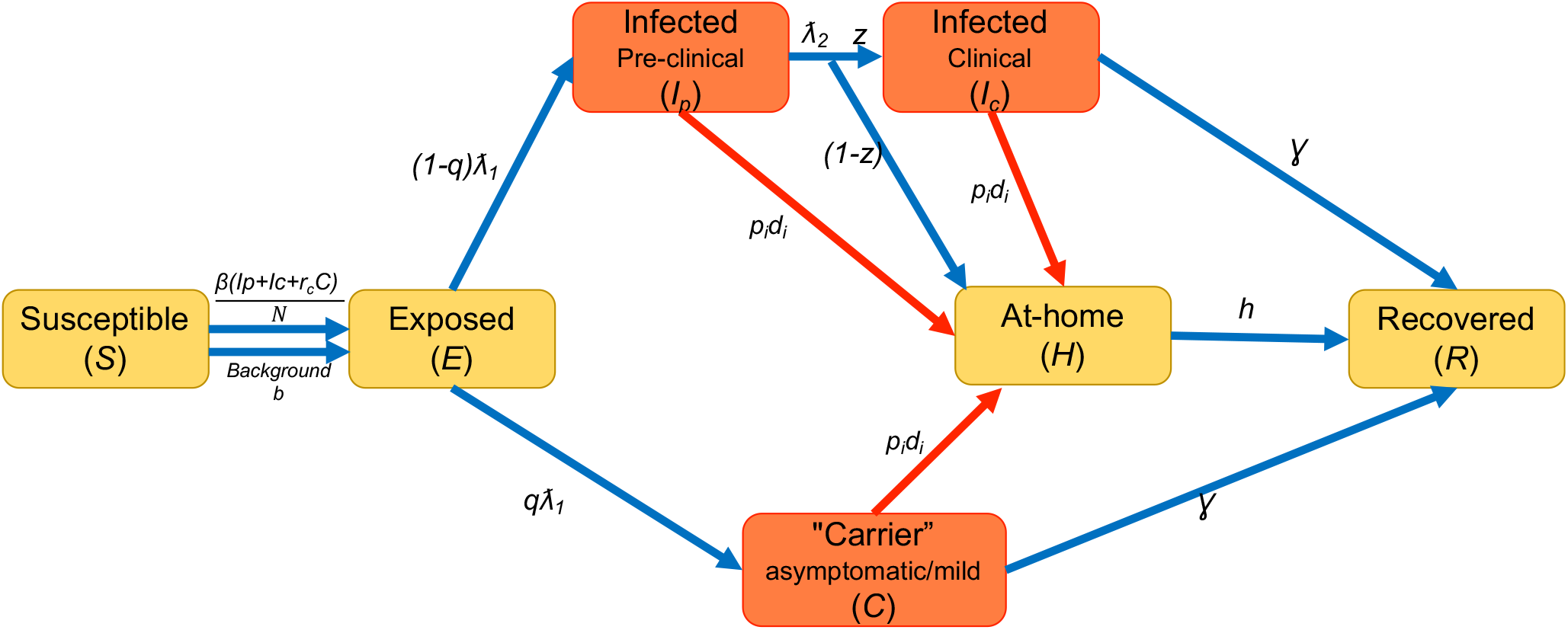
Schematic for compartmental transmission model. Orange boxes represent classes of individuals that contribute to transmission, and that are also detectable via PCR. Blue arrows represent processes that are modeled as differential equations in the deterministic model, and refer to transmission and disease progression processes. Red arrows represent screening and testing measures in the model, which occur at specific intervals. Red and blue arrows are included in the stochastic model. Parameters that determine the rate at which individuals transition between compartments are defined in Supplementary Table S1.

In both versions of the compartmental model, all individuals in the worker population are classified as belonging to a series of discrete compartments, and the number of individuals in each compartment at any given time is modeled through time (Figure 1). Compartments utilized for this model include: Susceptible (S)-Workers not yet infected; Exposed (E)-Workers that have contracted the virus but are still in the latent period of infection; proportion *q* of infected individuals become “carriers” (C)-workers whose infection is either truly asymptomatic or so mild that it is not detected; the remainder (1-*q*) of workers eventually become clinically infected (*I*_*c*_) but will pass through a pre-clinical asymptomatic phase (*I*_*p*_) prior to symptom onset; proportion *z* of clinically infected workers will continue to go to work while infected, but the remainder will stay at-home (*H*). After some length of time, infected individuals (*I*_*c*_ and *C*) and at-home individuals (*H*) will recover and return to work. Recovered individuals are assumed to be immune for at least the short time period of several months that is of interest here.

Parameters that determine the rate at which individuals transition between compartments are defined in Supplementary Table S1. Workers in the model can become infected by two means. First, transmission can occur from infected workers (*I*_*p*_, *I*_*c*_, and *C*) based on the effective reproduction number *R* for workplace transmission, with the asymptomatic class being proportionally (*R*_*c*_) less infectious than the clinical cases. Second, workers can become infected outside the workplace, which is represented by a constant background community transmission rate (*b*) and can be interpreted as a force of infection (per capita rate in which susceptible individuals become infected). Thus, the deterministic model is expressed a system of ordinary differential equations:

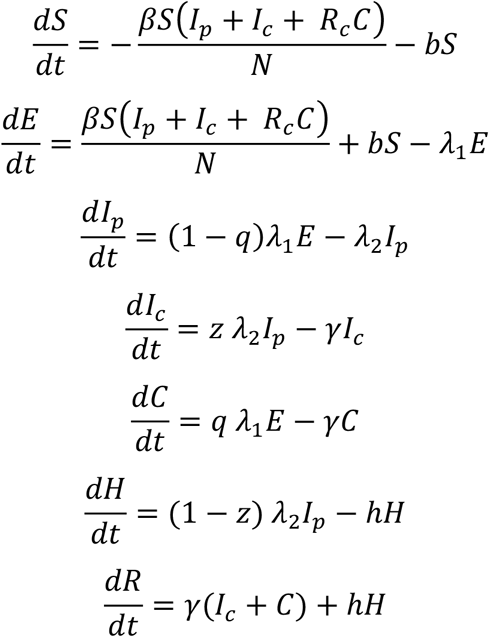

Where *λ*_*1*_, *λ*_*2*_ and *γ* were defined as the inverse of the latent, pre-clinical-infectious and clinical-infectious periods, respectively. *β*is the workplace transmission rate and is derived from *R* and the duration of infectiousness, and *h* is defined as the inverse of the period of time in which infected workers remain at home. Given the short duration of the time period of interest, the population size was considered constant and closed. At the onset of the outbreak, the population was considered fully susceptible with a single latently infected worker.

In the stochastic version of this model, we used the tau-leap method proposed by Gillespie (16) for incorporating stochasticity at each time step (17): each segment within the above equations is pulled from a Poisson distribution. For example, the number of workers leaving the exposed class was drawn from a Poisson distribution with mean (*λ*_1_ *E*). In addition, multiple testing/screening protocols were introduced to the model as events that occurred at specified intervals (red lines in Figure 1). Each testing/screening protocol *i* can be turned on or off in the model, and applied to a proportion of workers (*p*_*i*_) at a specified interval of days (*Freq*_*i*_). The testing/screening protocol detects infected workers with a certain probability based on the sensitivity (*se*_*i*_) of the method. Testing/screening either result in workers with positive tests to immediately go home (*delay*_*i*_ *=* 0 days), or after a user specified number of days due to delays in test results. Temperature-based screening was modeled to occur on a daily interval with no delays in results, but only detected clinically infected workers. PCR-based testing was modeled as being able to detect all workers in the *I*_*p*_, *I*_*c*_, or *C* classes, with a user specified delay in testing results.

An RShiny implementation of the stochastic version of this model was developed as is available at https://sumn.shinyapps.io/covidshcomp/. This modeling dashboard allows for customization of all parameter values and overlaps the trajectory of outbreaks based on user-defined testing/screening protocols.

This study was reviewed by the University of Minnesota Institutional Review Board (STUDY00012004), which made the determination that this research does not involve human subjects.

### Model calibration

We tuned the deterministic model to the epidemiological data (cumulative clinical cases and testing results) from each plant. Given the daily incidence data for three plants (Plants A – B) was based on self-reported PCR testing from private health care providers and one plant (Plant C) was based on ongoing company-provided testing of symptomatic workers, we assumed that workers who sought and received testing were experiencing clinical disease because testing was generally restricted to symptomatic patients at that time. Self-reported PCR-positive workers were also absent from work; therefore, we assumed the number of self-reported cases (Plants A and B) was equivalent to the at-home “H” class in our model. The percent of workers that were IgG-positive during company-initiated testing was assumed to be equivalent of the percent of workers in the *R* class at a particular point of time. Although it is possible for IgG to be detectable within two days of symptom onset (18), generally <70% of people had detectable IgG by 10 days of symptom onset (18, 19), while >85% had detectable IgG after 11-15 days (19-21). In our model, the length of time between infection and recovery was 11-15 days, which is why we believe that the percent of recovered individuals in the model is a reasonable approximation of the percent IgG-positive in the observed data.

Because most parameter values in the model are uncertain, we conducted a multivariate calibration exercise on uncertain parameters (Suppl. Table S1) using Latin hypercube sampling (LHS) and rejection sampling (22-24). We generated 10,000 parameter sets through sampling a Latin hypercube, which is an efficient method to sample multivariable parameter space. Model results were generated for each parameter set using the deterministic model. Parameter sets were then rejected if the modeled outbreak did not sufficiently resemble observed data, based on a set of criteria that was specific to each plant (cumulative number at-home, percent infected/PCR-positive or recovered/IgG-positive, see Supplementary Table S3 for criteria for each plant.). Goodness-of-fit criteria were based on epidemiological data from the earlier phases of plant-based outbreaks, up until the completion of company-initiated testing. Parameter value medians and interquartile ranges were summarized from candidate parameter sets that met the goodness-of-fit criteria. The median value was considered to be the most-likely value and used in subsequent model exploration.

Model fit was checked by plotting the observed epidemiological data against the model’s predictions. Here, 1000 simulations were performed with the stochastic model to estimate variability in model outcomes given the most-likely parameter values. For plants which experienced outbreaks of longer duration (B and D), it was apparent that although the calibrated parameter values produced simulated outbreaks that resembled the observed data in the early phase of the outbreak, the number of clinical cases was overestimated in the post-testing period. Therefore, we allowed *R* and *b* to be re-calibrated based on the post-testing data to account for altered disease dynamics, which may have emerged as a result of higher adherence to biosafety protocols (reduced *R*) or changes in disease prevalence in the community (reduced *b*).

We also performed a sensitivity analysis for the stochastic model using Latin hypercube sampling and random forest analyses, which is a common approach used for global sensitivity analyses in simulation modelling (22-25)(Supplementary text S2).

### Estimation of R

To further interrogate our estimate of *R* within the workforce population and to determine the sensitivity of *R* to the estimation method used, we estimated *R* using several accessory methods that use only data on the cumulative incidence of clinical cases, namely the exponential-growth (EG) method, maximum-likelihood (ML) method, and the time-dependent R (R-TD) method (26)(Supplementary text S3). The EG and ML method estimate *R* for the early phase of an outbreak (identified via built-in functions in the *R0* package as the period where cases approximated exponential growth). In contrast, time-dependent reproduction numbers (R-TD) allow *R* to be variable through time (26), and can be useful for monitoring epidemic trends, measuring the influence of interventions over time, and informing parameters for mathematical models (27). We used a procedure proposed by (15) to estimate effective R-TDs for each plant from the observed epidemic curves and generation time distribution. R-TD values were calculated on a daily interval and smoothed by weekly bins. 95% confidence intervals (CI) were obtained through simulation, as described elsewhere (26).

### Scenario testing

We evaluated the effectiveness of PCR-based screening (every 3, 7, 14, 28 days) in reducing the number of clinical cases relative to doing a temperature-screening only baseline. For each testing frequency, we also evaluated the effect of delays in testing results where infected workers remained at work until results were received (after 1, 3, or 5 days). We also evaluated the effectiveness of PCR-screening when 100% versus 75% of workers were tested, and across three population sizes (n = 100, 1000, and 2500 workers). Finally, we also evaluated each scenario if used proactively to prevent disease introduction, or reactively during the early, peak, post-outbreak stages. For the reactive scenarios, the starting number of individuals in each infection class was taken from the deterministic predictions (Early = first timepoint in which a cumulative of 0.5% of workers are clinically infected; Peak = timepoint with the highest number of infected individuals; Post = 60 days after the peak). We also considered an outbreak stage (35%-Imm.) in which testing is used to prevent re-introduction to an already partially immune population (35% of workers start as recovered to resemble the epidemiological situation of Plants B and D).

Based on the sensitivity analysis, it was apparent that model results were sensitive to the *R* and *b*, thus all scenarios were run for both high and low transmission parameters, yielding 1500 total scenarios. The choice of values for *R* (high=4, low=2) comes from a consensus of values from the above *R* estimations. The high background rate (*b* = 0.005) falls within the approximate range of background rates estimated across all plants. At the time of writing (November 15, 2020), daily incidence of >10 per 10,000 have been reported for Midwestern states, and assuming that there is a substantial underreporting, our high *b* is reasonable. The low background rage (*b* = 0.00015) is equivalent to ∼1.5 new cases per 10,000, which would be considered low to moderate at the time of writing and approximated background transmission in summer 2020 in the rural Midwest (28). Values for high/low *R* and *b* were compared to the results of the sensitivity analysis to ensure that the selected values spanned the variability in model outputs related to these parameters.

Other parameters were set to a single most-likely value based on either our model calibration or published literature (*R*_*c*_ = 0.75(29); Latent period = 5 days; Pre-clinical period = 1.5 days; Clinical period = 6 Days; proportion carriers = 0.5; proportion that go to work when clinically ill = 0.3; at-home period = 10 days; Temperature screening sensitivity = 0.7; PCR sensitivity = 0.9). Each scenario was simulated for 90 days, and 100 simulations were performed per scenario, yielding 150,000 total model runs.

### Software

Models were coded in R statistical software v3.6.0. Sensitivity analysis and model calibration were performed using the *tgp, randomForest*, and *randomforestexplainer* packages (30-32), and estimation of *R* was performed using the *R0* package (33).

## Results

### Descriptive epidemiology

Epidemic curves of the daily number and cumulative proportion of clinical cases plotted for each plant show that Plant C experienced the largest outbreak that plateaued at ∼25% of the workforce reporting clinical disease (PCR confirmed by company-sponsored testing available on an on-going basis, Figure 2a). Plant B experienced an outbreak that leveled off with just under 10% of the workforce reporting clinical disease (PCR confirmed by private health providers, Figure 2a), whereas Plant A experienced a much smaller outbreak of short duration. A list of mitigation measures employed by plants is listed in Table 2.

**Table 2.**
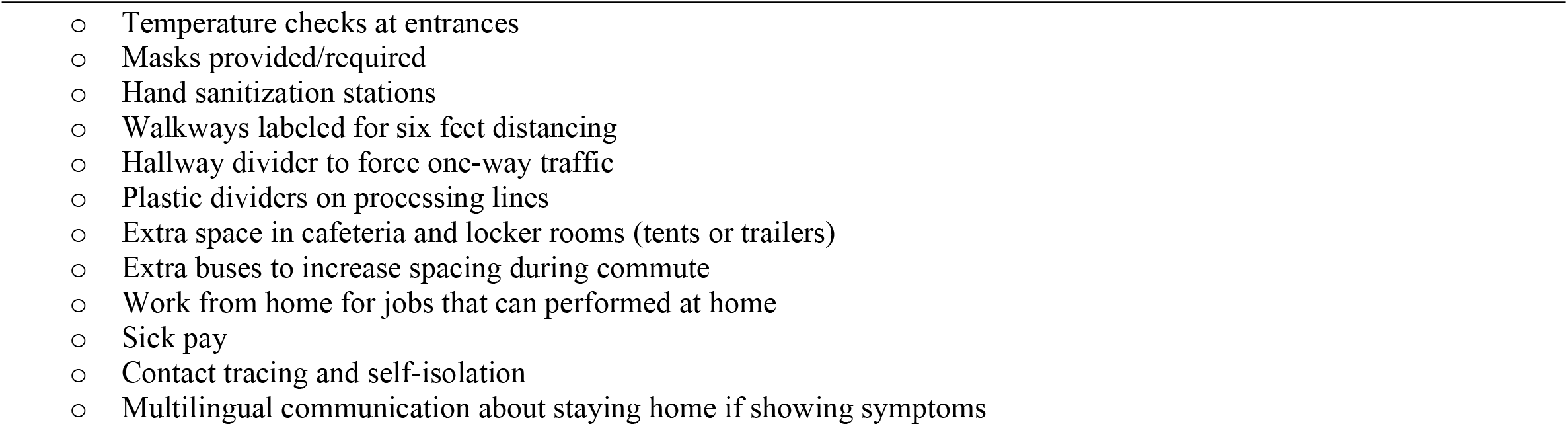
List of mitigation measures utilized by some or all plants

**Figure 2.**
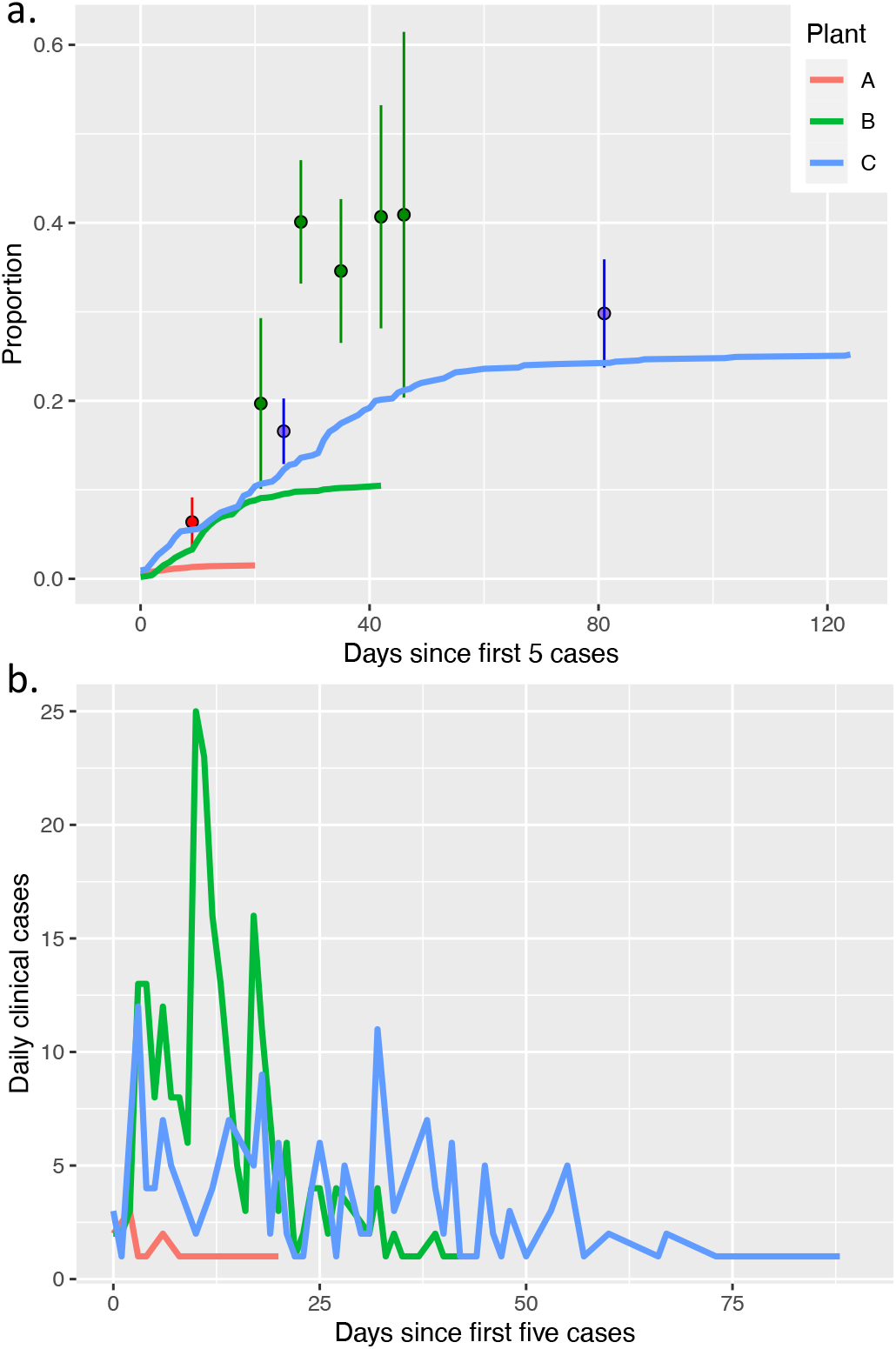
**a)** Cumulative clinical cases (at-home sick, confirmed PCR-positive) as a proportion of the worker population (lines) and proportion positive in company-initiated antibody testing (circles). b) Daily incidence of clinical cases.

For plant A, antibody testing was completed on May 1, 2020. At that time, 6.4% (95-CI: 2.6-9.2%) of workers were IgG-positive, 9.4% (95-CI: 6.1-12.7%, n = 298) were IgM-positive, and 15 workers (1.3%) were at-home sick/confirmed PCR positive (Table 1, Figure 2a). For Plant B, antibody testing was conducted over 4.5 weeks and was completed on May 12. No person was tested twice. By the conclusion of testing on May 12, a cumulative of 19.0% (95-CI: 15.4-22.5%) were IgM-positive, 35.8% (95-CI: 31.5-40.1%, n=472) were IgG-positive, and the cumulative number of workers at-home sick had leveled off at just over 200 workers (8.9%). However, the proportion of IgG-positive workers was 19.6% (10.1 −29.1%) in the first week of testing (April 17), and then increased and remained at ∼40% thereafter (weekly results shown in Supplement Table S2). Of all antibody tests performed, 3.3% (2.1-4.6%) were IgM(+)/IgG(-), 1.7% (0.8-2.6%) were IgM(-)/IgG(+), and 22.9% (20.0-25.9%) were IgM(+)/IgG(+). In Plant C, seroprevalence was 16.6% (12.9-20.3%) on May 19, at which point ∼90 workers at-home sick, and rose to 29.8% (23.7-35.9%) by July 14.

Antibody test results were compared against PCR results for Plant B workers (Supplementary Table S4). Of 42 workers that self-reported being previously PCR-positive (or presumptive positive), 35 (83%) were IgM- and IgG-positive. Additionally, of 419 workers who had never received a PCR-test, 123 (29%) were IgM- and IgG-positive, and 11 (2.6%) were IgM-positive and IgG-negative.

### Model calibration to outbreak data

The calibrated parameter values for each plant are summarized in Supplementary Table S1 and visually compared in Figure 3. The most likely values for parameters related to the progression of infection (e.g., duration of the latent period, pre-clinical period, and clinical period) were similar across plants, as was the proportion of workers estimated to go to work when experiencing clinical symptoms (most likely values ranging from 0.23 to 0.33). However, estimated *R*’s differed substantially among plants, as did background community transmission rate. Furthermore, a higher percentage of infections were estimated to be “carriers” at Plant A (∼80%) than at Plant B (∼50%) and Plant C (∼30%), and the relative infectiousness of “carriers” was estimated to be 0.26 – 0.46.

**Figure 3.**
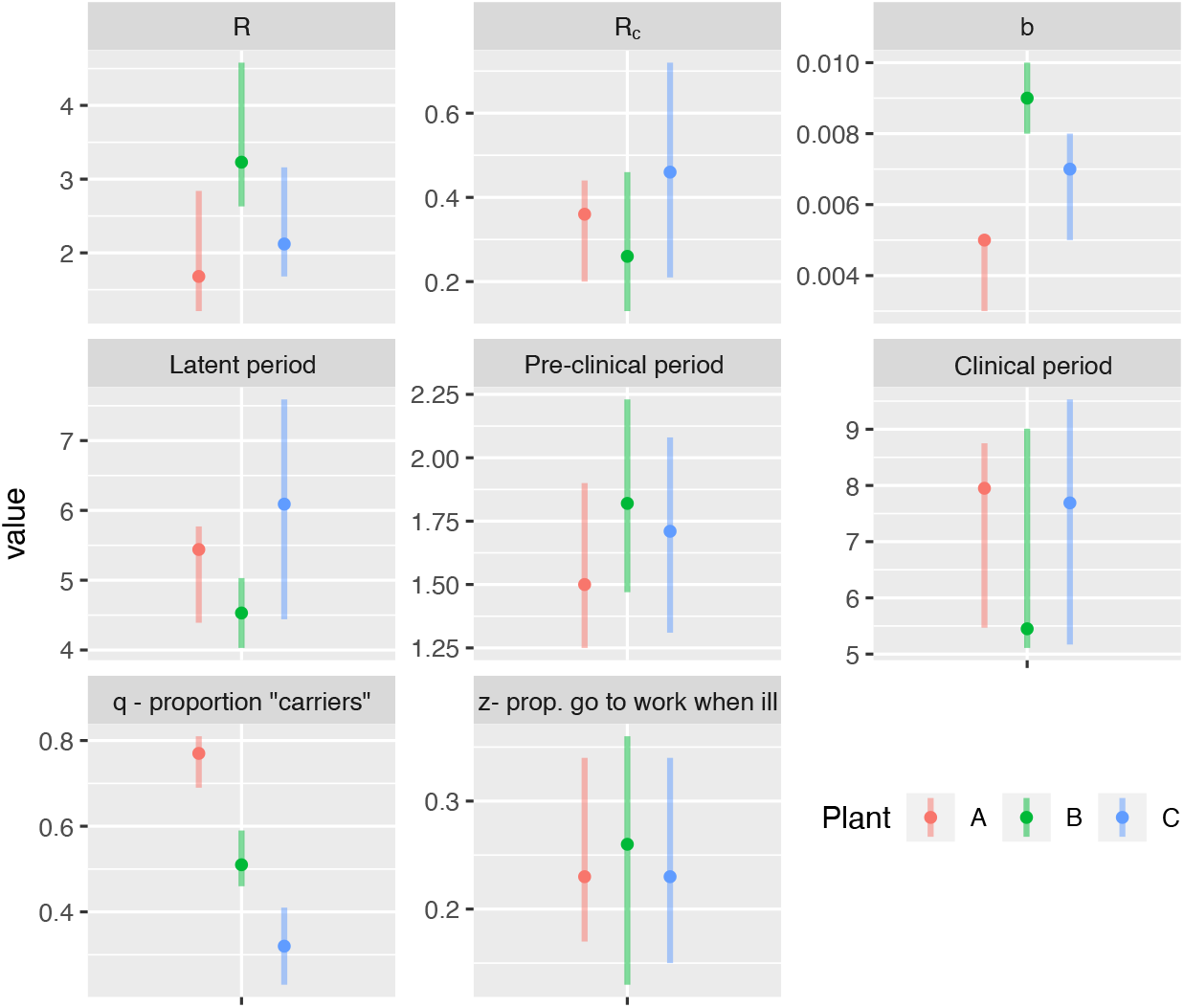
Comparison of the most-likely value and interquartile range of each parameter value in the tuned models.

The model fit was evaluated against the observed number of cumulative clinical infections. The model fit reasonably well earlier in the outbreak, which were the data used for model tuning, but the number of cases were overestimated in the latter phase of the outbreaks (Supplementary Figure S2). Therefore, the parameter values of *R* and *b* were re-calibrated for the post-testing phase of the outbreaks for Plant B and D (Plant A had very few additional cases post-testing). For Plant B, the adjusted *R* estimate for the post-testing period (after the first week of testing) was 0.81 (0.64-1.07), and *b* dropped to 0.001 (0.0006 – 0.002). For plant C, *R* was reduced by 60% post-testing. The final stochastic-runs of the models are shown in Figure 4. The number of transmission events due to transmission in workplace (driven by *R*) versus community (driven by *b*) is show in in supplementary Figure S3.

**Figure 4.**
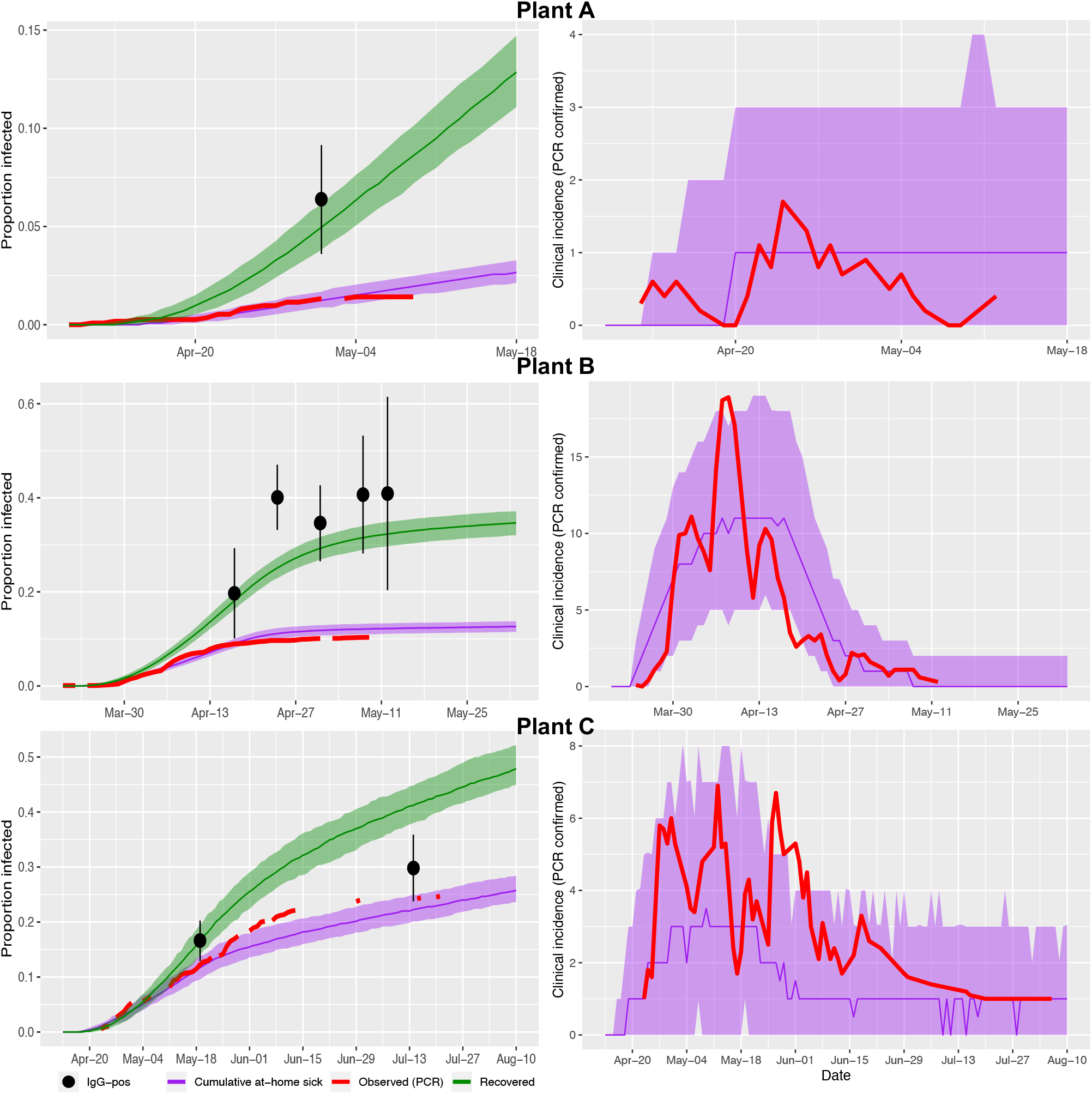
Output of stochastic models for plants that experienced large outbreaks (Plants B and D), showing (left) the cumulative proportion at-home sick or recovered, and (right) the smoothed daily incidence of clinical cases (4-day moving average). For both cumulative proportions and incidence plots, observed clinical case data are shown in red. Model predictions are based on the median (colored lines) and 50% prediction interval (shaded area) summarized from 1000 runs of the stochastic model. Black dots indicate the observed proportion IgG-positive, with whiskers showing standard error of estimates.

Based on a sensitivity analysis of the stochastic model, the parameters that were most associated with variation in the cumulative number of clinical cases were the transmission rate parameters (background transmission rate, *R*, and *R*_*c*_, Figure 5a). Cumulative clinical cases decreased with higher *q* (proportion of infections that are “carriers”, Supplementary Figure S4). Given the fundamental importance of *R* and *b*, we used the random forest analysis to visualize the interaction between these two variables (Figure 5b).

**Figure 5.**
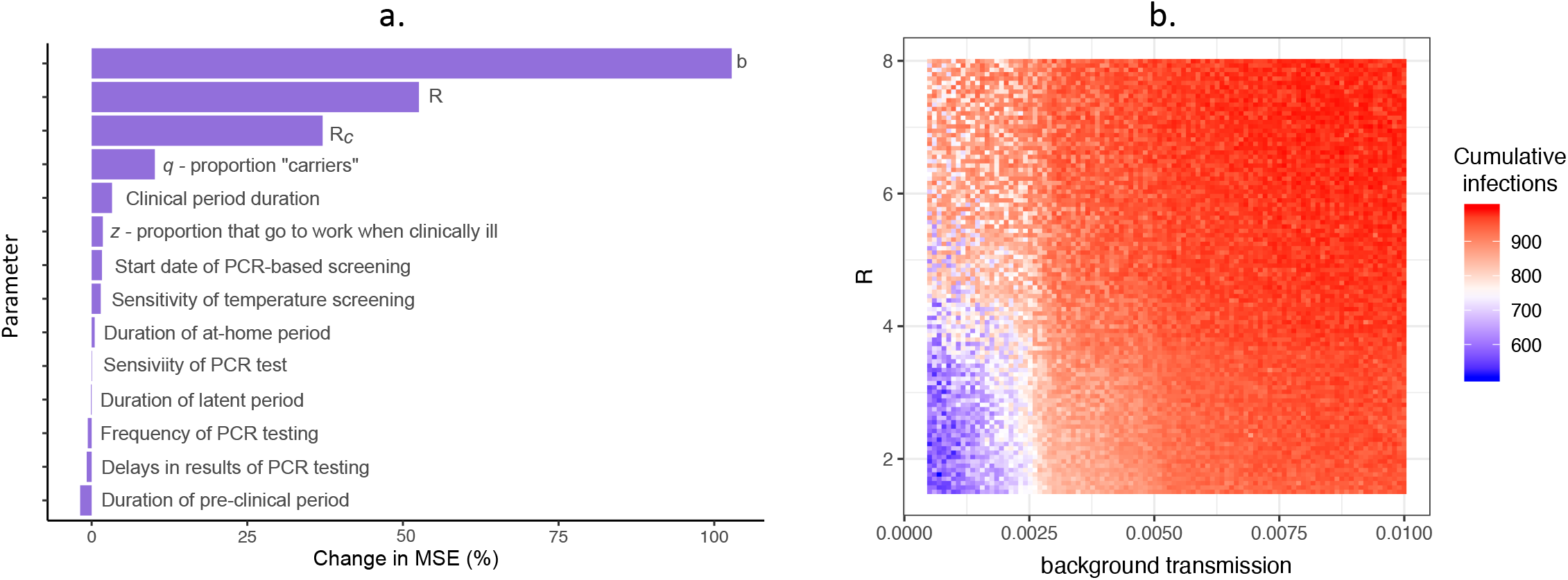
a) Variable importance plot for random forest sensitivity analysis, with most important parameters ranked from highest to lowest in terms of their association with the predicted number of cases. Percent change in MSE was calculated as the percent change in mean squared error when the explanatory variable was permuted relative to the outcome. b) Predicted cumulative infections from the random forest sensitivity analysis based on the interaction between *R* and background transmission rate. Shading represents the predicted cumulative number of infections for a given value of *R* and *b* (background transmission).

### Estimation of R

In addition to calibration of the compartmental model, we used three alternate methods to estimate *R*. Across all four methods, *R* ranged from 1.7 – 2.7 for Plant A, 3.0 – 4.4 for Plant B, and 2.1 – 3.2 for Plant C (Figure 6a, Supplementary Table S5). Based on this analysis, *R* = 2 was considered “low,” and *R* = 4 was considered “high” in subsequent scenario testing. Across all plants, the time-dependent *R*’s initially were >2 for the first one to two weeks, and then fell to below or close to 1 by week two or three (Figure 6b). Results of these three methods are influenced by generation time assumptions. Here, we show the results assuming a generation time of six days, as that is most compatible with our calibrated parameter estimate of ∼5 days for the duration of the latent period (Table 2). However, similar results were obtained when assuming a generation time of four days (Supplementary Table S5 and Figures S6-7).

**Figure 6.**
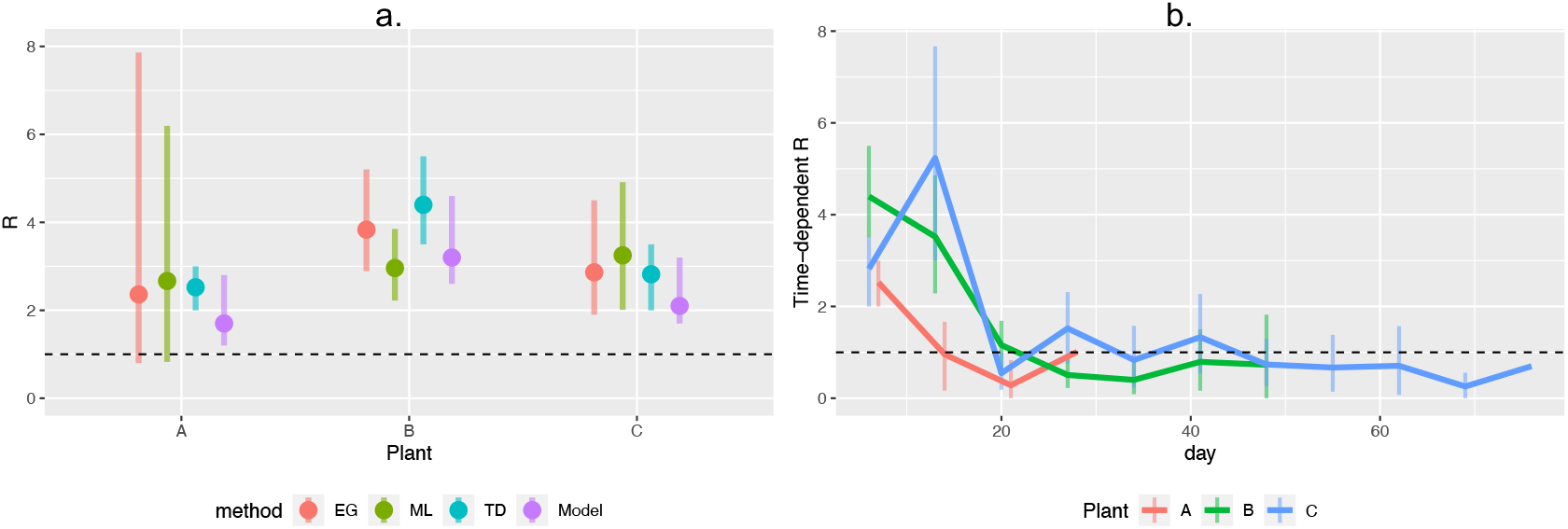
**a)** Comparison of R estimates using different estimation techniques. Colors indicate each technique: EG: Exponential growth; ML: Maximum likelihood; TD: R-TD during the first week; Model: Estimate from the tuned compartmental model. b) Time-dependent R estimated at a weekly interval for each plant. In both panels, black hashed line shows R = 1.

### Scenario testing

To evaluate the effectiveness of different PCR screening protocols, we modeled how the number of workplace infections was influenced by the frequency of full workforce PCR-testing (every 3, 7, 14, or 28 days) and delays in test results (1, 3, or 5 days) across three population sizes (n = 100, 1000, and 2500). We also explored scenarios in which screening began at five different outbreak stages (prevention, early, peak, post, and 35% immune). Due to model sensitivity to *R* and *b*, each scenario was modeled with a factorial combination of high/low *R* (*R* = 2 / 4) and *b* (b = 0.00015 / 0.005). For each 90 day scenario, we summarized the cumulative number of additional infections (only infections that occurred after the onset of PCR screening were counted), cumulative number of additional clinical cases, proportion of infections that were due to workplace transmission, number of infections detected by PCR (Supplementary Figures S7-11), the number and proportion of clinical cases averted (relative to a temperature-screening only baseline, Figure 7). All analyses were repeated with only 75% of workers undergoing testing, and results were largely similar (Supplementary Figures S12-16).

**Figure 7.**
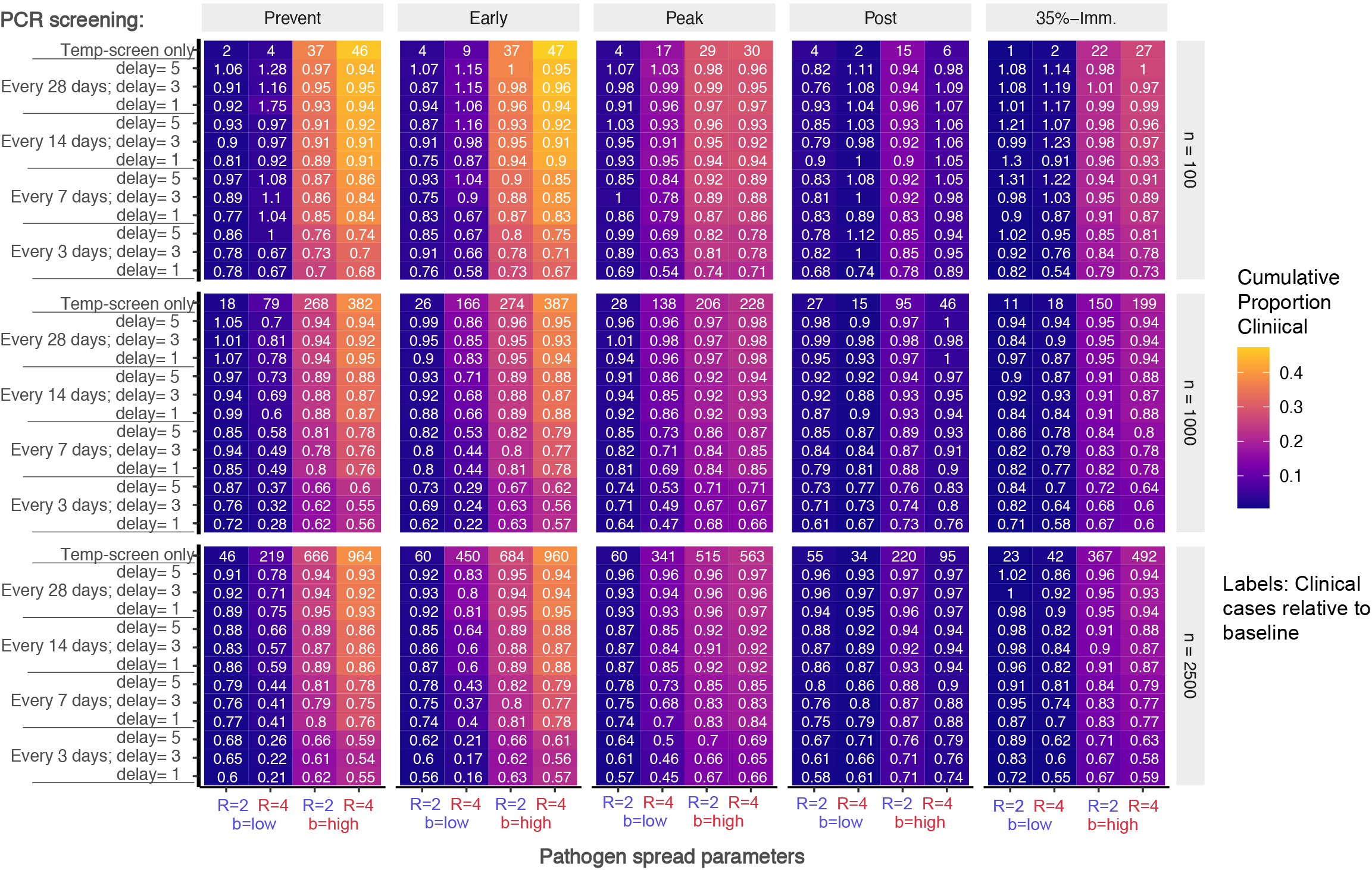
Proportion of expected clinical cases averted by PCR-screening relative to baseline of temperature screening alone. Results are stratified according to stage at which PCR-screening is implemented (Prevention, Early, Peak, Post, 35% Immune), workforce size (n = 100, 1000, 2500), and high/low workforce *R* and background transmission rates. Absolute number of expected clinical cases under the baseline is labeled in the top line of each colored block, and each cell label shows the clinical cases relative to baseline (*Cases*^*scenario*^ */ Cases*^*baseline*^). Colored shading indicates the cumulative proportion of the workforce that experience clinical disease.

## Discussion

In this study, we described the epidemiological situation in three pork processing facilities in the United States, including characterizing each plant’s epidemic curves and results of company-initiated cross-sectional sampling. We then developed a mathematical model, calibrated to each plant’s data, that accounted for asymptomatic, pre-symptomatic, and background “community” transmission. All plants experienced SARS-CoV2 infections in their workforce beginning in March and April 2020, a time period in which SARS-CoV2 was rapidly spreading throughout the country and scientific understanding of its epidemiology, transmission, and appropriate protective and mitigation measures was rapidly evolving. The epidemic curves of two of three plants demonstrated relatively large outbreaks, with ∼10% to 25% of workers reporting PCR-confirmed clinical disease. Daily increases in cases in the workforce plateaued after approximately two and six weeks for Plants B and D, respectively. The parameter values estimated during model calibration revealed both similarities and differences across plants. These differences are more apparent for the three plants experiencing larger outbreaks, and may have been related to variability in plant location, case identification methods, and potentially implementation and compliance with biosafety measures.

Plant B, for example, was located in a region where high levels of community transmission early in the pandemic may have contributed to the steep epidemic curve. A greater role of community spread within this region was supported by model calibration results for Plant B, in which the estimated background transmission rate was notably higher than in other plants. During model calibration of Plant B, it was difficult to find parameter combinations in which the cumulative clinical cases (at-home/PCR confirmed) remained below the observed 10%, while also achieving ∼40% recovered (assumed to be IgG+ positive). It is possible that the self-reported clinical cases (at-home sick / PCR-positive) underestimated the true number of clinical cases. Only cases that were PCR-confirmed were included, and some people may have been unable to get tested even though they were showing mild symptoms. Compared to the other plants, this plant’s outbreak began in mid-March, a time period when testing was limited, many were unaware of COVID-19 clinical signs, and symptoms could have been confused with other seasonal illnesses. Alternatively, the discrepancy between cumulative clinical cases and percent of the workforce recovered may have emerged because we assumed that IgG-positive individuals were exclusively found in the recovered class. In the model, the mean time from exposure to recovered and assumed IgG-positive would be ∼10-12 days. However, it is possible for individuals to have detectable IgG as early as 4 days post-infection, though >10 days would be more likely (19). If some individuals have detectable IgG prior to recovery (10-12 days), then some non-recovered individuals would contribute to the IgG prevalence. This could explain why observed IgG prevalence was higher than the predicted proportion of people in the recovered class.

Plant C was the only plant to offer company-sponsored PCR-testing for diagnostic purposes on a rolling basis. Although testing was generally restricted to those showing clinical signs, including very mild clinical signs, the ease of access to PCR-testing may have led to more cases being PCR-confirmed than other plants, and thus an apparent larger outbreak with a cumulative of ∼25% of workers clinically affected. Compared to other plants, the observed epidemic curve based on PCR-confirmed clinical cases was much more closely aligned to the antibody prevalence based on cross-sectional surveillance, suggesting that fewer cases went undocumented. This is also reflected in the much lower estimated values of *q* (proportion of infections that are asymptomatic). Plant C also had a policy that asked all household and carpool contacts of a potential case to self-isolate at the same time as the employee showing clinical symptoms, i.e. they all isolated from first report of clinical symptoms, not when test result is received. These contacts were only tested if clinical signs developed within the 14-day quarantine period. Thus, when illness started in these contacts, they were already outside of the workplace, and their contact tracing led to few or no new workplace contacts. If they developed clinical signs while in quarantine but tested negative, they were still not released from quarantine before the end of the 14-day period. It is notable that the sero-positivity in this population was only slightly lower than that of Plant B, and Plant B’s clinical curve leveled off at just 10%. These observations suggest potential under-documentation of clinical cases at Plant B, or a wider clinical definition of what would warrant administration of a PCR test at Plant C. Plant C offered PCR testing as needed on site if at least one clinical sign of COVID-19 was present, however mild, but testing was generally not offered to asymptomatic employees, even those in quarantine due to limits in test availability at the time. Thus, it is difficult to determine whether Plant C truly experienced a larger outbreak, or simply a better documentation of cases.

With the above considerations in mind, there are several general insights from the model that warrant further discussion. The majority of *R* estimates across plants fell between ∼2.5 to 3.5, regardless of estimation method, indicating that each infection resulted in around three additional infections during the outbreaks’ early period. Interestingly, although this is a high-density population, this estimate is not markedly different from the values estimated for the general population (34-36). In addition, an analysis of *R* through time showed that *R* declined to around 1 within two to three weeks, suggesting that outbreaks were brought under control fairly rapidly. For example, Plant C experienced a decrease in cases at the end of April, corresponding to the date that screening questions were implemented at the point of pick-up (rather than at the plant’s entrance) for those employees who take a bus from a nearby community. Plant C also experienced a spike in cases in early May, which corresponded to an outbreak at a neighboring meat processing plant and the discovery that employees from both plants sometimes share a household. The pre-shift screening questions were adjusted to ask about shared living arrangements or close contact with an employee from the neighboring plant over the previous 3 days, this resulted in a significant number of employees being asked to self-isolate (with pay) for the 14-day quarantine period. Other measures employed at plants include staggered shifts to minimize overlap in workers, barriers and signage to enforce social distancing, extra buses hired to increase spacing during commute, providing masks and hand-sanitizer, and contact tracing combined with self-isolating employees who showed any clinical symptoms or whose household members were experiencing clinical signs or being tested for COVID-19 (Table 2). While some of these measures were in place in plants even before substantial workplace cases were observed, discussions with company personnel indicate that compliance with biosafety measures may have increased after the company-initiated cross-sectional sampling was performed due to greater awareness of the scope of the outbreak. The dynamic nature of *R* also highlights the importance of not simulating results too far into the future, as underlying assumptions about transmission rates and other model parameters may change through time.

Interestingly, our results also suggest a potential role of population immunity in slowing infections, though we do not advocate that this should be a purposeful strategy for disease mitigation. If the initial value of *R* is assumed to be 3, then the critical immunization threshold to achieve herd immunity would be 66% (1 – 1/*R*_*0*_). However, if biosafety measures reduce by *R* by one half (*R* = 1.5), this shifts the threshold to 33%. Interestingly, this is not too different from the antibody prevalence reported in Plants B and D. If herd immunity is indeed playing a role in these plants, then it would be expected that the worker population no longer can sustain a large-scale outbreak, though there still may be a consistent low level of cases due to exposure in the community. Indeed, Plant C showed a decrease in new cases during July, even though cases reported in the general community were increasing at this time. In addition, none of the plants have experienced a “second wave” as of November 2020, and case counts at the plant align well with what would be expected from current incidence levels in surrounding communities. That being said, worker turnover and uncertainties about the duration of immunity both could change the percent of the population that is antibody positive through time. Additionally, if worker contact rates change and/or compliance with biosafety measures relaxes, this would allow *R* to shift back upward and consequently the critical immunization threshold needed for herd immunity would also shift upward.

One of the most challenging aspects of controlling SARS-CoV2 is the presence of asymptomatic carriers that are capable of transmitting the virus. In this project, the word “carrier” was loosely applied to those testing PCR or antibody positive, but who reported no symptoms. However, there was not adequate documentation or follow-up to definitely conclude that they were asymptomatic. Thus, the “carrier” compartment in the model more accurately represented both true asymptomatic infections as well as those with mild symptoms that either were not aware of infection or were unable to obtain a test. It is possible that the estimated proportion of infections that were “asymptomatic” was lower in Plant C (∼30%) because company-sponsored PCR-testing was available on a rolling basis. The 50% asymptomatic rate in Plant B (located in a regional epicenter) may be because more workers sought and received testing, compared to Plants A that were distant from epicenters at the time (as discussed above, Plant C’s on-site testing may have resulted in fewer undetected cases). Plant A had a modeled asymptomatic rate of >75%, which matches reasonably with the cross-sectional testing data in which 93% of antibody-positive workers at Plant A reported no symptoms.

Community transmission is key to understanding and modeling the unfolding of outbreaks at plants. Here, we chose to handle community transmission as a constant background rate. As described above, it appears that plant-to-plant variability in estimated background rates reflected observed regional trends, with Plant B estimated to have the highest background rate during this period. However, we did not allow for the background rate to change dynamically through time. Also, while we can compare our background transmission rates to CDC county-level incidence data, it is also important to note that many workers tend to socialize and live together with other workers both from the same plant and other plants, sometimes as a part of immigrant communities. Thus, the community transmission experienced by workers is likely not completely decoupled from the transmission dynamics within the workplace, nor is it reflective of dynamics in the general population. For these reasons, worker exposure to SARS-CoV2 in the community may be higher than the general public due to such heterogeneities in community contacts. It is also important to note that the nature of our data limits the conclusions we can draw about community transmission, given that comparable data from communities were not available. Thus, our estimates of community transmission should be interpreted cautiously.

With this in mind, a general insight from our model indicates the importance of community exposure is dependent on the stage of the outbreak. Community spread accounted for more transmission events than workplace transmission in the very early and post-peak periods of the simulations. However, once a workplace outbreak was established, workplace transmission accounted for the majority of transmission events (Supplementary Figure S3). Community transmission appeared to kickstart workplace outbreaks, resulting in the outbreak reaching a “tipping point” of exponential growth more quickly. Indeed, our sensitivity analysis suggests that there is a background transmission threshold of ∼25 cases per 10,000 people, above which workplace outbreaks are distinctly larger (Figure 5).

Finally, we used the calibrated model to evaluate the effectiveness of routine PCR-screening at reducing disease circulation within a plant across a range of scenarios (Figure 7, Supplementary Figures S7-16). As expected, more cumulative clinical cases occurred at higher values *R* and *b*, though the differences between high and low *b* were much larger than the differences between high and low *R* (color of shading in Figure 7). Indeed, in a population of 1000, the baseline expected number of additional clinical cases (prevention stage) was 18 and 17 for the low and high *R* scenarios when *b* was low, and 268 and 382 for low and high *R* when *b* was high.

Across all outbreak stages and population sizes, the proportion of clinical cases averted (cell labels in Figure 7) was most influenced by testing frequency and less influenced by delays in results, *R, b*, and the proportion of workers tested. Testing every three days generally averted about 25% to 40% of clinical cases, while testing every seven days averted somewhere between 13% and 20% of clinical cases. Testing every 14 and 28 days typically averted 7 to 13% and 2 to 8% of clinical cases, respectively. However, it should be noted that the number of additional clinical cases expected was sometimes quite small (top number in each block in Figure 7), particularly for population sizes of 100. Thus, the absolute number of cases averted should be considered alongside other factors. It is also notable that even frequent testing may not prevent a large outbreak when background transmission is high and population level immunity is low (prevention and early outbreak stages).

Our results are similar to other modeling studies examining routine PCR screening for SARS-CoV2 (14). For example, testing every 2 days was deemed necessary to limit cases to controllable levels at college campuses (37). Other models also emphasize the importance of biweekly testing (11, 14), with the predicted reductions in transmission related to the assumed value of *R*, other control measures employed (11), and delays in testing results (14). Our results also mirror others in that testing frequency was found to be more important than test sensitivity (14, 37). Notably, in (14), the relative number of cases under each testing scenario were not strongly influenced by use of an agent-based versus homogenous mixing modeling framework, which supports the use of a homogenous mixing model for our smaller-scale workplace model.

In conclusion, we summarized SARS-CoV2 outbreaks experienced in three pork processing plants during the spring of 2020. Through calibrating a mathematical model to the epidemiological data from these plants, we demonstrated that *R* was generally between 2 and 4 in the early stages of these outbreaks, but rapidly declined within two to three weeks. This slowing in transmission is clearly evident in the epidemic curves of each plant, and is most likely a consequence of implementation and adherence to biosafety measures employed at each plant potentially in combination with population immunity. We found substantial heterogeneity across plants in asymptomatic rates and the relative infectiousness of “carriers,” but this variation is more likely an artifact of awareness, access to tests, and differences in reporting as opposed to differences in biology. Finally, routine PCR-screening was shown to reduce the number of clinical cases in the workforce, but the absolute number of cases averted depended on assumptions about disease transmissibility and the stage of the outbreak. Even frequent testing may not prevent a large outbreak within the workforce. While the model was calibrated to meat processing plants, the structure of the model and insights about testing are generalizable to any workplace where large number of people work in close proximity.

## Data Availability

Data in this study are confidential.

## Acknowledgements

We are grateful to companies that shared their data, as well as company representatives that contributed to the development of the model and interpretation of results. We also thank members of the University of Minnesota Swine Group for their contributions to this work early on in the process. This work was funded by the Swine Disease Eradication Center.

## Notes

### Competing Interest Statement

The authors have declared no competing interest.

### Funding Statement

This work was funded by the Swine Disease Eradication Center at the University of Minnesota

