## Supplementary materials for "Modeling transmission dynamics and effectiveness of worker screening programs for SARS-CoV-2 in pork processing plants"

***Supplementary text S1. Incidence data***

For Plants A and B, individual confirmed cases were dated to either the test result date or the absentee date, whichever was earliest. These two dates were a median of 0 days apart (IQR: -0.75 – 2 days). For Plant C, tests were administered on the first day of symptoms, and all close contacts home-quarantined (with full pay) at the same time.

***Supplementary text S2. Sensitivity Analysis***

We performed a sensitivity analysis for the stochastic model using Latin hypercube sampling and random forest analyses, which is a common approach used for global sensitivity analyses in simulation modelling [1-4]. Random forest is an ensemble machine learning method that is based on the consensus of hundreds of randomized decision trees [5]. Unlike regression approaches, random forests can handle more complex data with numerous interacting and multicollinear variables with non-linear effects on the outcome [6,7], making them a highly suitable approach for understanding the marginal contribution of variation in each parameter on the outcome after controlling for the effects of all other parameters. Here, the sensitivity analysis not only included the rate parameters from the deterministic model, but also parameters that control the frequency, diagnostic sensitivity, and delays in results for company-initiated PCR testing and daily temperature screening (Table 2). Briefly, we generated 1,000 parameter sets sampled from a Latin hypercube, and simulated 1,000 epidemics for each set (population size=1000, duration of simulation = 365 days). The median number of cumulative infections was summarized for each set, and this value was used as the outcome in the random forest. Parameter values were included in the random forest as explanatory variables to explain variation in the outcome [1,8]. To assess the overall performance of the model, the percent of variance explained by the random forest was calculated. Variable importance was assessed by calculating the percent increase in mean standard error (MSE) of predictions when x is permuted relative to y, with larger changes in MSE indicating that the variable is an important predictor of the outcome. Partial dependence plots were constructed to assess the marginal effect of model parameters on model outputs after accounting for the effects of all other parameters.

***Supplementary text S3. Accessory methods for estimating R***

For the EG method, *R* was inferred from the early phase of the outbreak, during which time the epidemic curve approximates exponential growth. The EG method combines the observed per capita change in number of new cases per day with an assumed generation time to estimate the population's *R* [9,10]. Second, we applied the maximum-likelihood (ML) method proposed in [9,11], which is built on the assumption that the number of secondary cases for each infected individual follows a Poisson distribution with a mean *R* that can be estimated though a likelihood-based approach. Generation time was assumed to follow a gamma distribution with a mean of six days and standard deviation of one day [12-14]. The "early phase" time period (identified via built-in functions in the *R0* package as the period where cases approximated exponential growth) used for the EG and ML methods is shown in Supplementary Figure S2.

**Supplementary Table S1.** Parameter definitions, symbols and types. Plausible range indicates the reasonable range of values for each parameter. (*) indicate parameters that were only used for the stochastic model sensitivity analysis, whereas non-starred parameters were used both for deterministic model calibration and stochastic sensitivity analysis. Calibrated values show the most likely (median) values from model calibration for each plant's data, with the interquartile range shown in parenthesis.

| Definition | | Type | Plausible range | Plant A | Plant B | Plant C |
|---|---|---|---|---|---|---|
| Reproduction number | $R$ | disease | 1.0 – 8.0 [15-25] | 1.7 (1.2 – 2.8) | 3.2 (2.6 – 4.6) | 2.1 (1.7 - 3.2) |
| Relative infectious of carriers | $R_c$ | disease | 0.05 – 1.0 [26-31] | 0.36 (0.22-0.4) | 0.26 (0.13-0.46) | 0.46 (0.21 – 0.72) |
| background transmission rate(b) | $b$ | disease | 0.0001- 0.01 | 0.005 (0.003-0.005) | 0.009 (0.008-0.01) | 0.007 (0.005 – 0.008) |
| Percent asymptomatic carriers (q) | $q$ | disease | 0.1 - .95 [12,26,30-36] | 0.77 (.69-.81) | 0.51 (0.46 – 0.59) | 0.32 (0.23 – 0.41) |
| Latent period | $1/\lambda_1$ | disease | 3 – 9 [17,37,38] | 5.4 (4.3 – 5.7) | 4.5 (4.0 – 5.0) | 6.1 (4.4 – 7.6) |
| Pre-symptomatic period | $1/\lambda_2$ | disease | 1-2.5 [13,39] | 1.5 (1.3 – 1.9) | 1.8 (1.5 – 2.2) | 1.7 (1.3 – 2.1) |
| Clinical (infectious) period | $1/\gamma$ | disease | 2.5 – 11 [13] | 7.95 (5.5 – 8.8) | 5.5 (5.1 – 9.0) | 7.7 (5.2 – 9.5) |
| Prop. people that go to work when ill | $z$ | Worker behavior | .05 – 0.5 [40] | 0.23 (0.17 – 0.34) | 0.26 (0.13 – 0.36) | 0.23 (0.15 – 0.34) |
| At-home isolation period (default = 10 days) | $1/h$ | Worker behavior | 7 – 21* | | | |
| Sensitivity of daily temperature screening (detection of $I_c$) | $d_{temp}$ | Temp. screening | 0.6 – 0.9* [41,42] | | | |
| Sensitivity of PCR testing (detection of $I_c$, $I_c$, $C$) | $d_{pcr}$ | PCR testing | 0.85 – 1.0* [43,44] | | | |
| Frequency of company-initiated PCR testing (interval in days) | | PCR testing | 7 – 28* | | | |
| Start day of company-initiated PCR testing | | PCR testing | 1 – 183* | | | |
| Delay in results for PCR test | | PCR testing | 0 – 7* | | | |

**Supplementary Table S2.** Results of antibody testing at Plant B.

| Date | IgM-IgG | No. tested | Prop. positive | 95-CI: Lower bound | 95-CI: Upper bound |
|---|---|---|---|---|---|
| **4/17/20** | Neg-neg | 66 | 0.75757576 | 0.65418418 | 0.86096734 |
| **4/17/20** | Neg-pos | 66 | 0.01515152 | -0.0143196 | 0.04462265 |
| **4/17/20** | Pos-neg | 66 | 0.04545455 | -0.0047995 | 0.0957086 |
| **4/17/20** | Pos-pos | 66 | 0.18181818 | 0.08876576 | 0.2748706 |
| **4/17/20** | IgG-pos | 66 | 0.1969697 | 0.10101864 | 0.29292076 |
| **4/24/20** | Neg-neg | 192 | 0.546875 | 0.47646108 | 0.61728892 |
| **4/24/20** | Neg-pos | 192 | 0.015625 | -0.0019177 | 0.03316767 |
| **4/24/20** | Pos-neg | 192 | 0.05208333 | 0.02065366 | 0.08351301 |
| **4/24/20** | Pos-pos | 192 | 0.38541667 | 0.31657346 | 0.45425987 |
| **4/24/20** | IgG-pos | 192 | 0.40104167 | 0.33171529 | 0.47036805 |
| **5/1/20** | Neg-neg | 133 | 0.65413534 | 0.57329704 | 0.73497363 |
| **5/1/20** | Neg-pos | 133 | 0.02255639 | -0.0026791 | 0.04779183 |
| **5/1/20** | Pos-neg | 133 | 0 | 0 | 0 |
| **5/1/20** | Pos-pos | 133 | 0.32330827 | 0.24381432 | 0.40280222 |
| **5/1/20** | IgG-pos | 133 | 0.34586466 | 0.26502637 | 0.42670296 |
| **5/8/20** | Neg-neg | 59 | 0.57627119 | 0.45017919 | 0.70236318 |
| **5/8/20** | Neg-pos | 59 | 0.01694915 | -0.0159885 | 0.04988676 |
| **5/8/20** | Pos-neg | 59 | 0.01694915 | -0.0159885 | 0.04988676 |
| **5/8/20** | Pos-pos | 59 | 0.38983051 | 0.26538099 | 0.51428003 |
| **5/8/20** | IgG-pos | 59 | 0.40677966 | 0.28143158 | 0.53212774 |
| **5/12/20** | Neg-neg | 22 | 0.59090909 | 0.38545491 | 0.79636327 |
| **5/12/20** | Pos-pos | 22 | 0.40909091 | 0.20363673 | 0.61454509 |
| **5/12/20** | IgG-pos | 22 | 0.40909091 | 0.20363673 | 0.61454509 |

**Supplementary Table S3.** Key indicators from observed data, goodness-of-fit criteria used for rejection sampling, and the number of parameter sets that met criteria and were used to define the calibrated parameter values.

| | A | B | C |
|---|---|---|---|
| Key indicators from observed data | • Time.10H: Day in which a cumulative of 10+ people were at-home sick (i.e., self-reports), must be within 18 days of first case<br>• Time.15H: Day in which a cumulative of 15+ people were at-home sick, which was observed to occur 6 days after Time.10H (4-12 days)<br>• Proportion recovered at Time.15H is in the 95-CI of the IgG-positive rate: 3.6 - 9.2% | • Time.20H: Day in which a cumulative of 20+ people were at-home sick (i.e., self-reports)<br>• Time.100H: Day in which a cumulative of 100+ people were at-home sick, which was observed to occur 7 days after Time.20H<br>• Time.200H: Day in which a cumulative of 200+ people were at-home sick, which was observed to occur 9 days after Time.100H<br>• Time.20R: Day in which 20% of workers are recovered (presumed IgG+), observed to occur same week as 200H<br>• R.3wk_post20: Percent recovered two weeks post time.20R, which was observed to be 40±13% | • Time.5H: Day in which cumulative of 5+ people were at-home sick<br>• Time.50H: Day in which cumulative of 50+ people were at-home sick, observed to be 14 days (±4 from time.5H)<br>• Time100.H: Day in which cumulative of 50+ people were at-home sick, observed to be 14 days from time.50H (±4 days)<br>• Time.90H: Day in which cumulative of 90+ people were at-home sick<br>• R.at.time.90H: Percent recovered (presumed AB+) was observed to be 16.6% (12.9-20.3%) at time.90H |
| "Goodness-of-fit" criteria | • Time.10H < 18<br>• Time.10H+4 days < Time.15H < Time.10H + 12<br>• 3.6% < R at Time.15H < 9.2% | • 2 < time.100H < 10<br>• Time.100H+5 < time.200H < Time.100H + 14<br>• Time.20R < time 200H +5<br>• 27% < R.3wks_post20 < 53% | • Time.5H +10 < time.50H < time.5H+18<br>• Time.50H + 10 < time.100H <- time.50H + 18<br>• 12.9% < R.at.time.90H < 20.3$ |
| Parameter sets meeting criteria | 18/10,000 | 17/10,000 | 255/20,000 |

**Supplementary table S4.** Number of Plant B workers positive and negative to IgG and IgM according to self-reported PCR status.

| | IgM/IgG | | | | |
|---|---|---|---|---|---|
| **Self-reported PCR test:** | **-/-** | **-/+** | **+/-** | **+/+** | **Total** |
| Not Tested | 278 | 7 | 11 | 123 | 419 |
| Negative COVID19 test | 8 | 0 | 0 | 3 | 11 |
| Positive COVID19 test | 2 | 1 | 3 | 28 | 34 |
| Presumptive Positive | 1 | 0 | 0 | 7 | 8 |
| **Total** | **289** | **8** | **14** | **161** | **472** |

**Supplementary Table S5.** Estimated *R'*s for Plants A-C based on four methods.

| Method | A | B | C |
|---|---|---|---|
| Exponential growth (EG) | 2.3 (0.8 – 7.8) | 3.8 (2.9 – 5.2) | 2.8 (1.9 – 4.5) |
| Maximum-likelihood (ML) | 2.7 (0.8 – 6.2) | 3.0 (2.2 – 3.8) | 3.2 (2.0 – 4.9) |
| Time-dependent R (TD-R, week 1) | 2.5 (2.0 – 3.0) | 4.4 (3.5 – 5.5) | 2.8 (2.0 – 3.5) |
| Model | 1.7 (1.2 – 2.8) | 3.2 (2.6 – 4.6) | 2.1 (1.7 – 3.2) |

**Supplementary Figure S1.** The time period during which the epidemic curve best approximated exponential growth is shown between the blue-hashed lines for Plants A, B, and C. This period was identified through the built-in default settings utilized by the *R0* package.

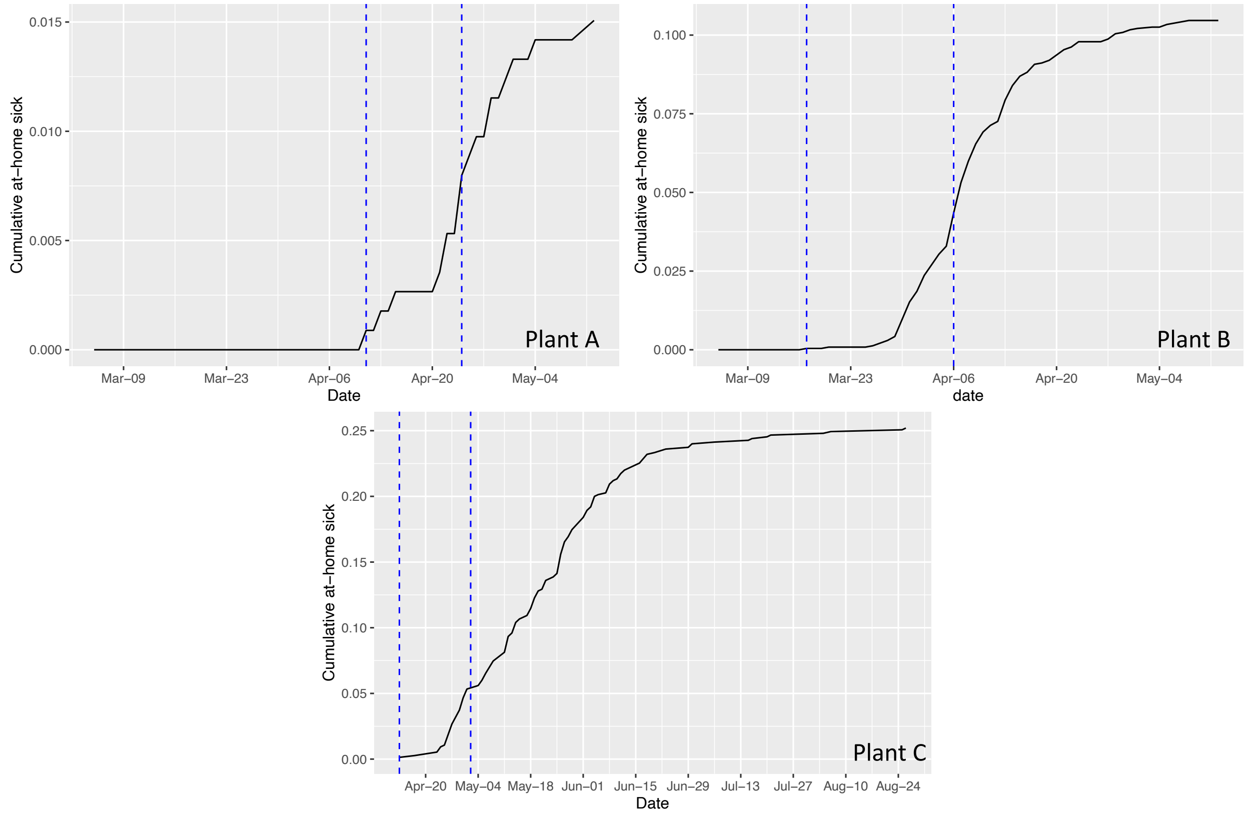

**Supplementary Figure S2.** Top: Cumulative proportion of at-home sick workers predicted by the tuned models (black) compared to the observed incidence data (red). Bottom: Cumulative proportion infected predicted by the tuned models (black line) compared to the observed antibody testing data (circles). Data prior the blue dashed line (date of conclusion of company-initiated testing) were used for initial model.

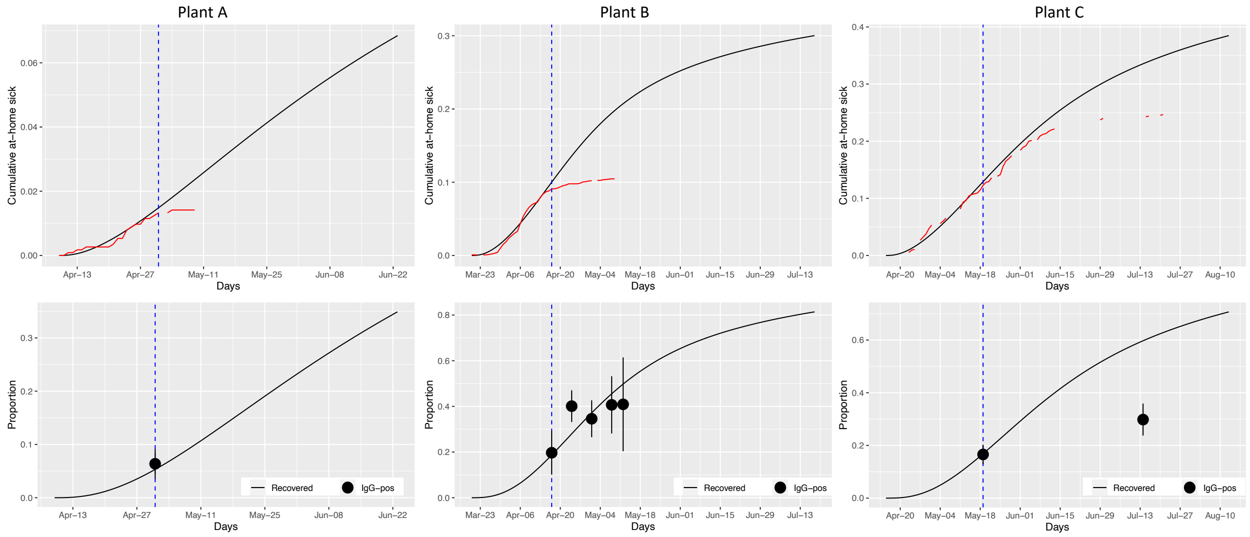

**Supplementary Figure S3.** Number of transmission events due to workplace-based transmission (red) versus community transmission (black).

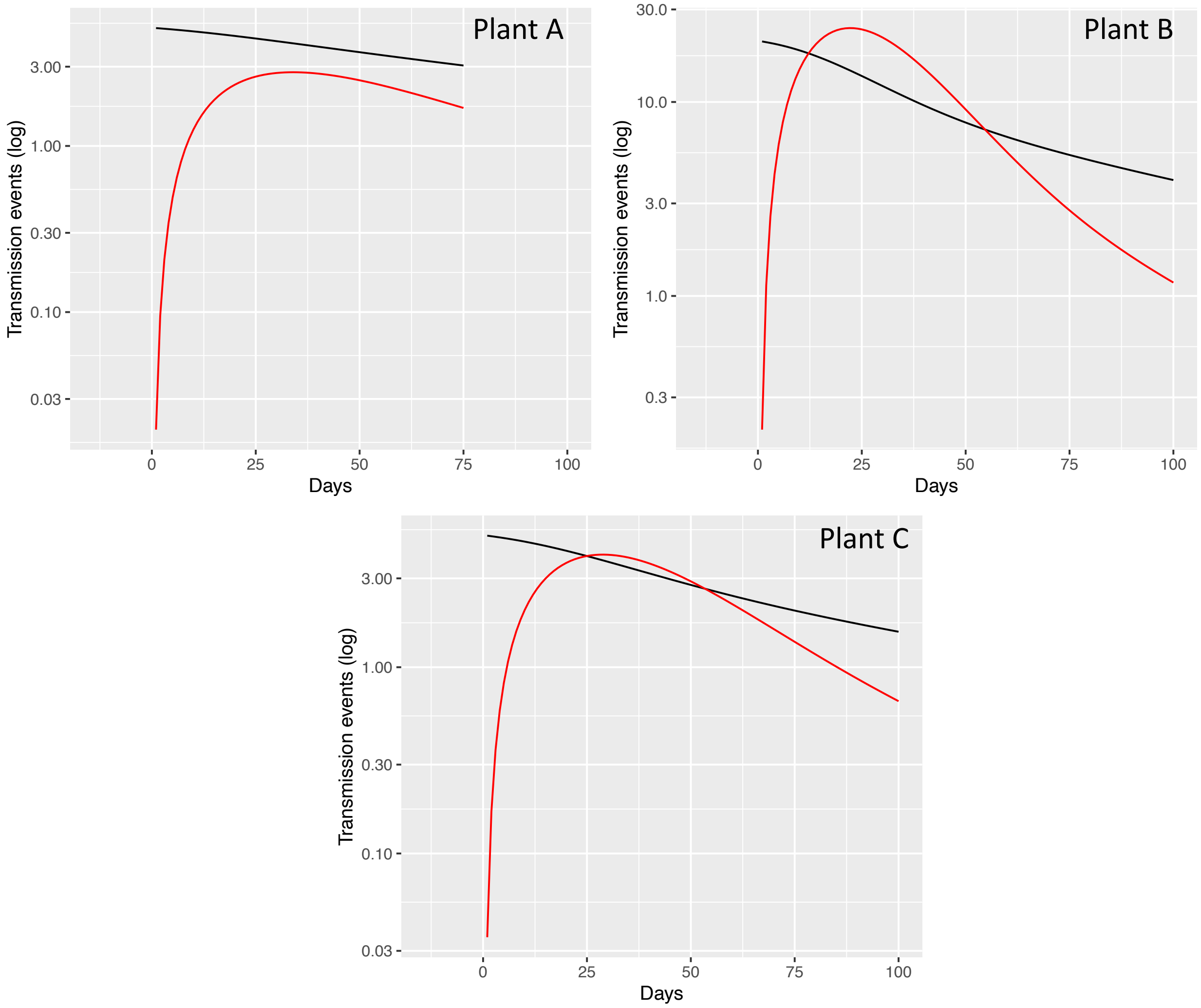

**Supplementary Figure S4.** Partial dependence plots showing the marginal effect of varying each parameter on the cumulative number of infections.

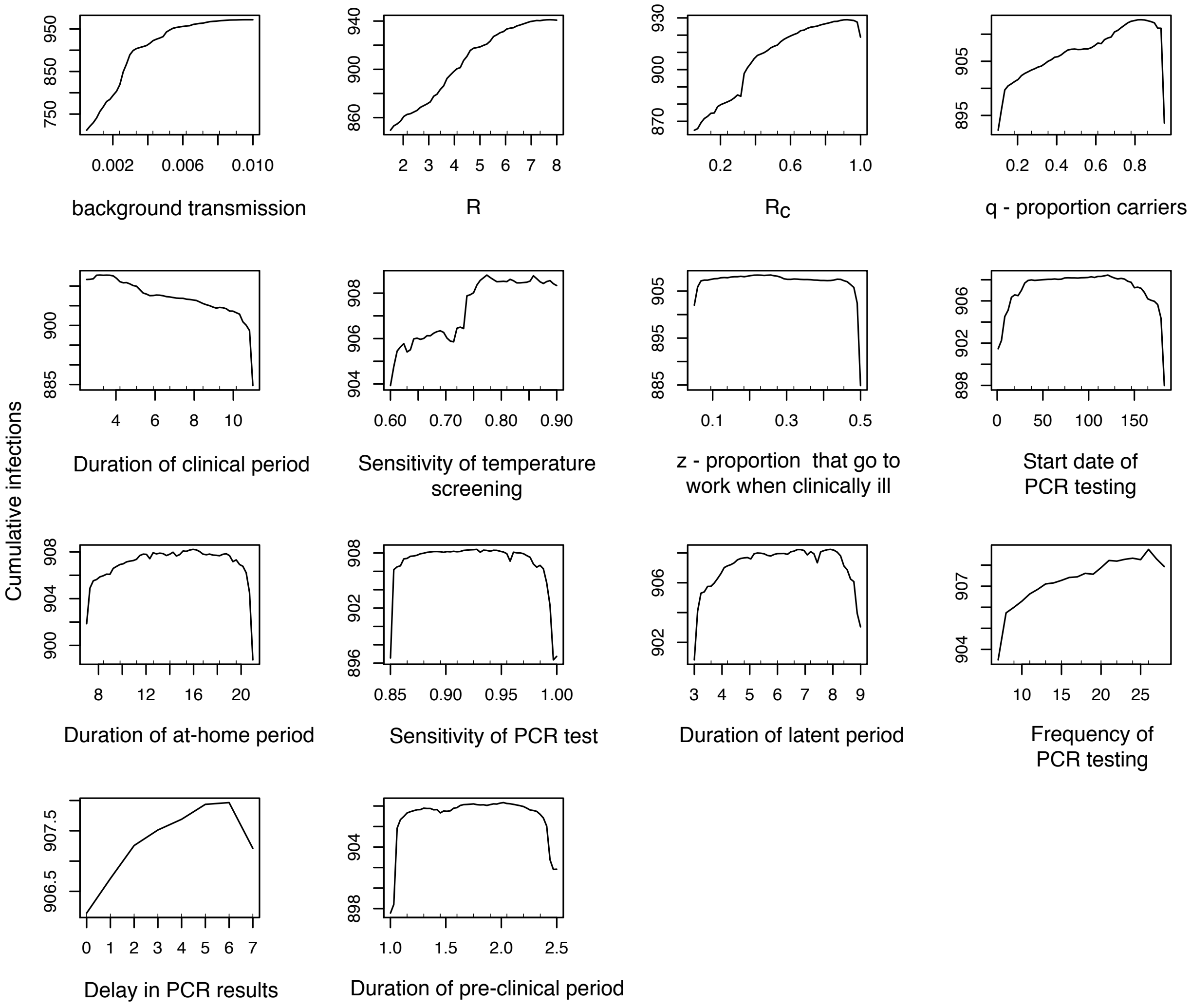

**Supplementary Figure S5.** Comparison of R estimates using different estimation techniques. Colors indicate each technique: EG: Exponential growth; ML: Maximum likelihood; TD: R-TD during the first week; Model: Estimate from the tuned compartmental model. Shape indicates the assumed generation time.

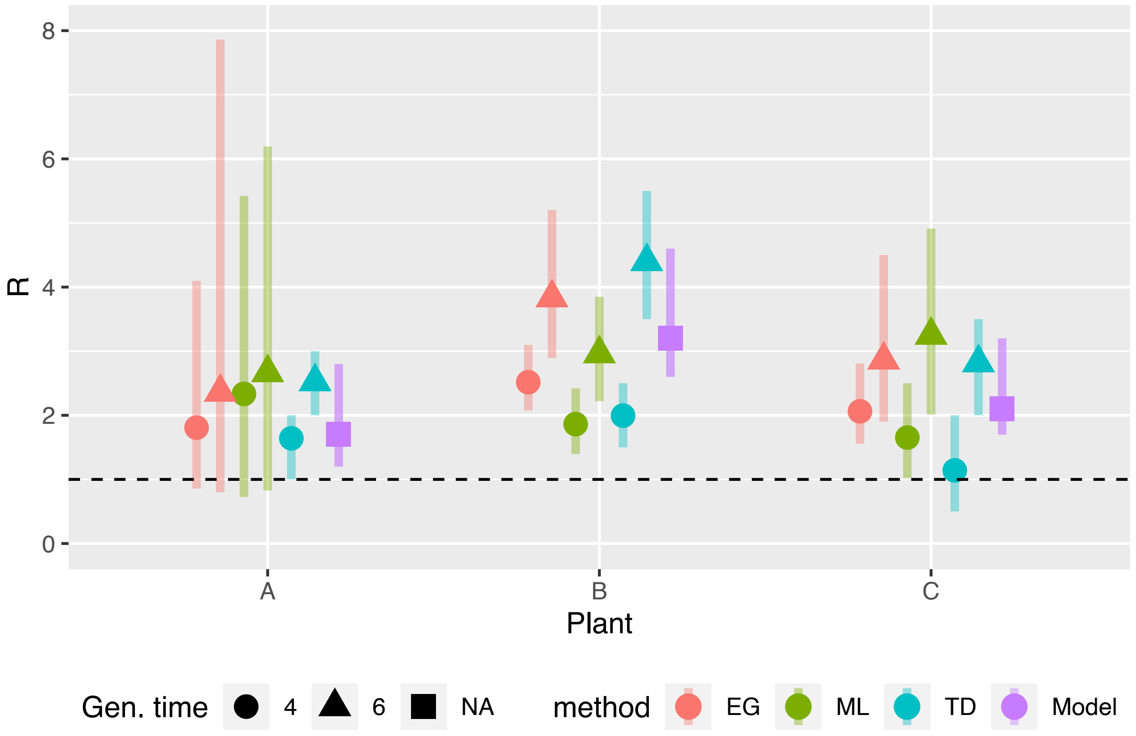

**Supplementary Figure S6.** Time-dependent R estimated at a weekly interval for each plant, assuming generation time of four days (top) and six days (bottom).

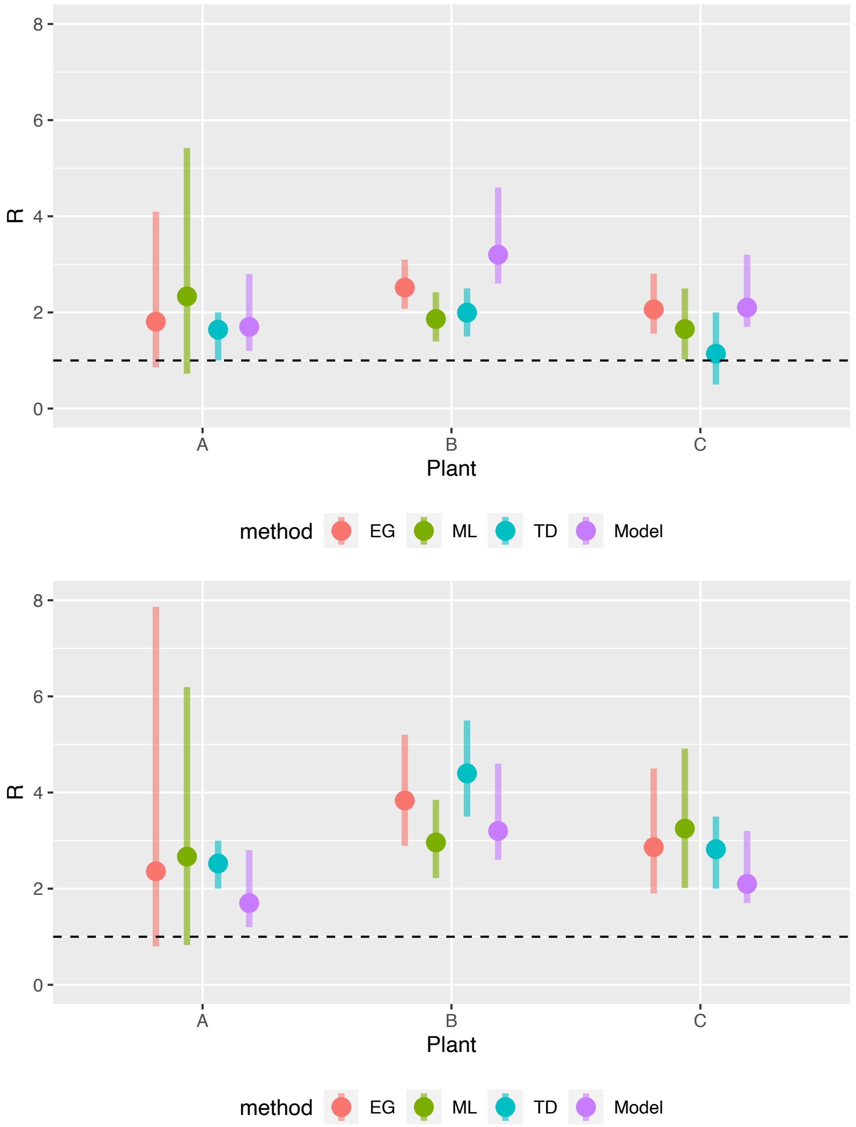

**Supplementary Figure S7.** Cumulative number of additional infections. Results are stratified according to stage at which PCR-screening is implemented (Prevention, Early, Peak, Post, 35% Immune), workforce size (n = 100, 1000, 2500), and high/low workforce *R* and background transmission rates. Each cell label shows the absolute number of additional infections. Colored shading indicates the cumulative proportion of the workforce that is infected.

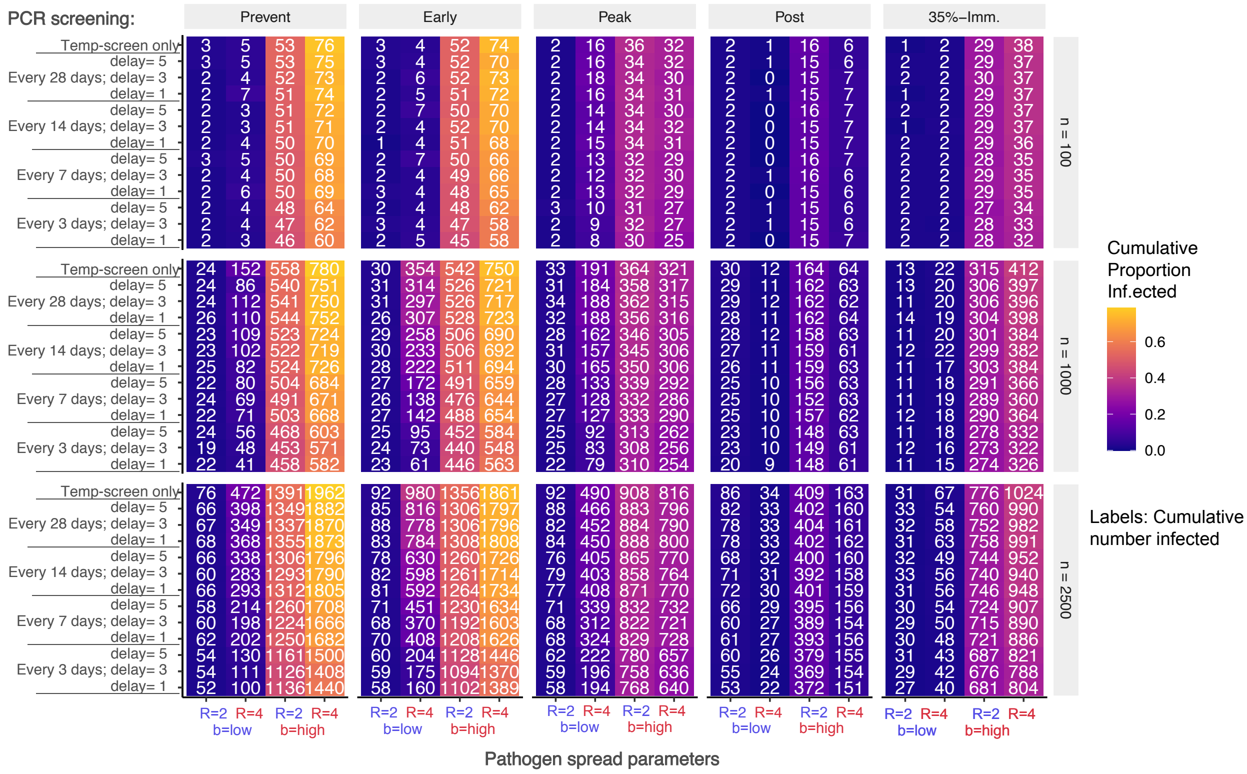

**Supplementary Figure S8.** Cumulative number of additional clinical cases. Results are stratified according to stage at which PCR-screening is implemented (Prevention, Early, Peak, Post, 35% Immune), workforce size (n = 100, 1000, 2500), and high/low workforce *R* and background transmission rates. Each cell label shows the absolute number of additional clinical cases. Colored shading indicates the cumulative proportion of the workforce that experiences clinical disease.

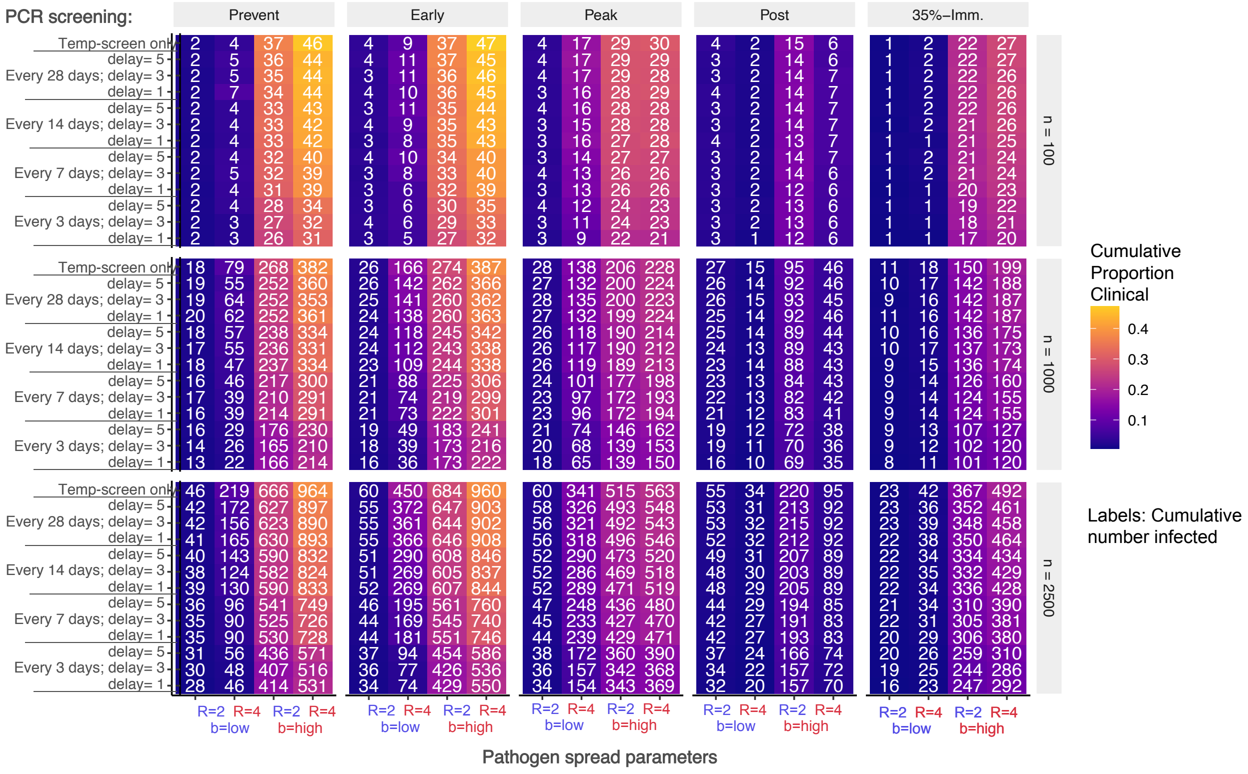

**Supplementary Figure S9.** Proportion of infections due to workplace transmission. Results are stratified according to stage at which PCR-screening is implemented (Prevention, Early, Peak, Post, 35% Immune), workforce size (n = 100, 1000, 2500), and high/low workforce *R* and background transmission rates. Each cell label shows the cumulative number of additional workplace transmission events. Colored shading indicates the proportion of all transmission events that were attributed to the workplace.

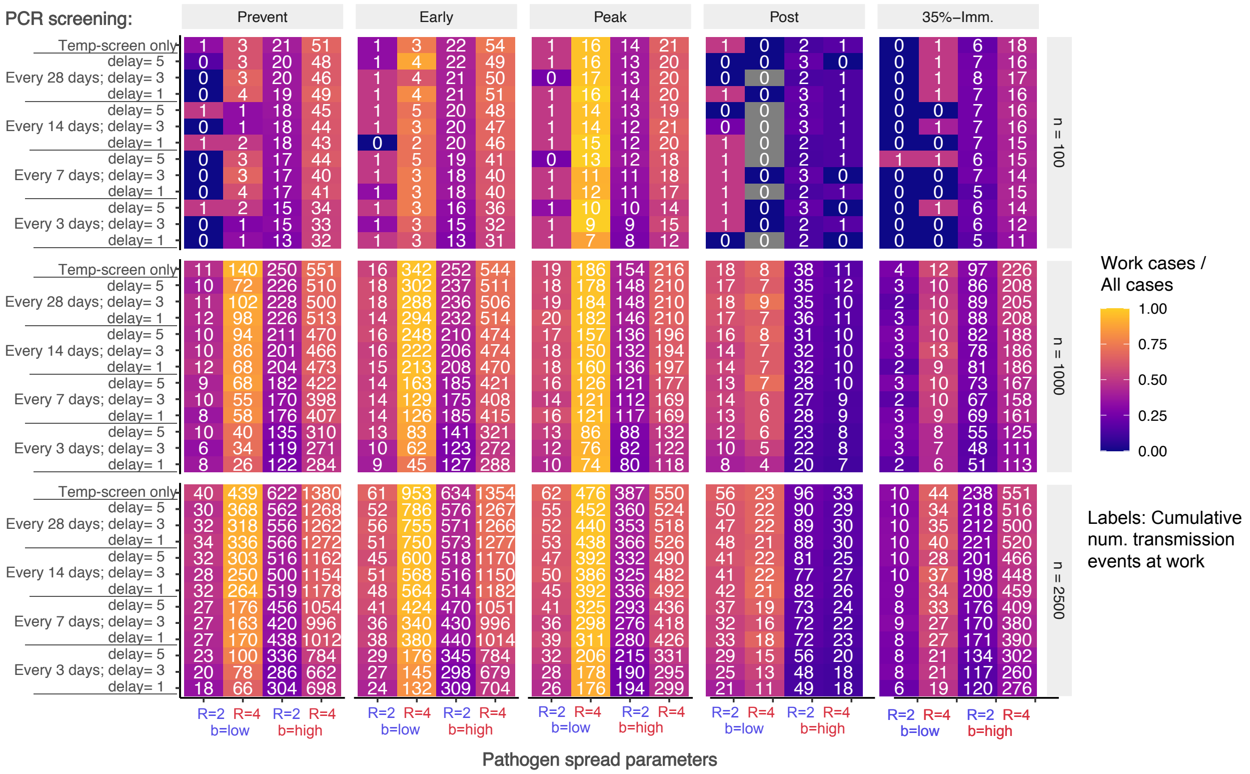

**Supplementary Figure S10.** Number of infections detected via PCR-screening. Results are stratified according to stage at which PCR-screening is implemented (Prevention, Early, Peak, Post, 35% Immune), workforce size (n = 100, 1000, 2500), and high/low workforce *R* and background transmission rates. Each cell label shows the absolute number of infections detected via PCR-screening. Colored shading indicates the proportion of infections that were detected.

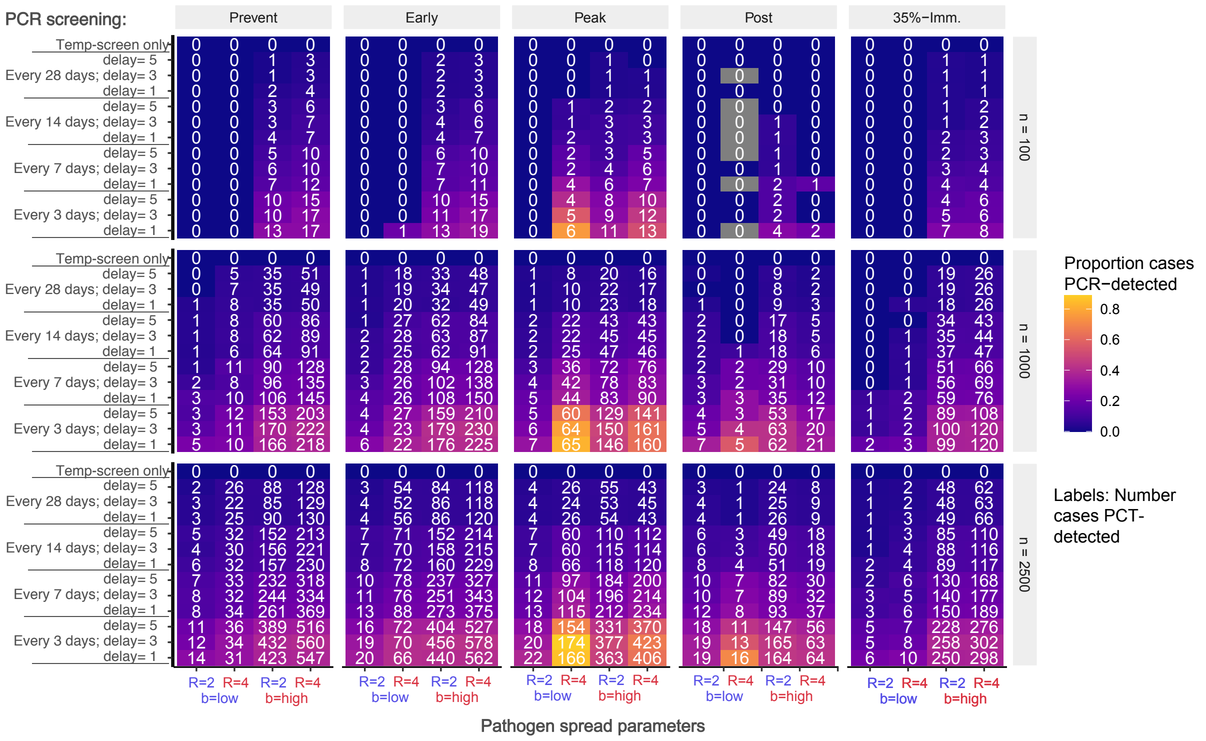

**Supplementary Figure S11.** Number of expected clinical cases averted by PCR-screening relative to baseline of temperature screening alone. Results are stratified according to stage at which PCR-screening is implemented (Prevention, Early, Peak, Post, 35% Immune), workforce size (n = 100, 1000, 2500), and high/low workforce *R* and background transmisison rates. Each cell label shows the absolute number of infections averted relative to the baseline (top row of each cell block). Colored shading indicates the number of cases averted.

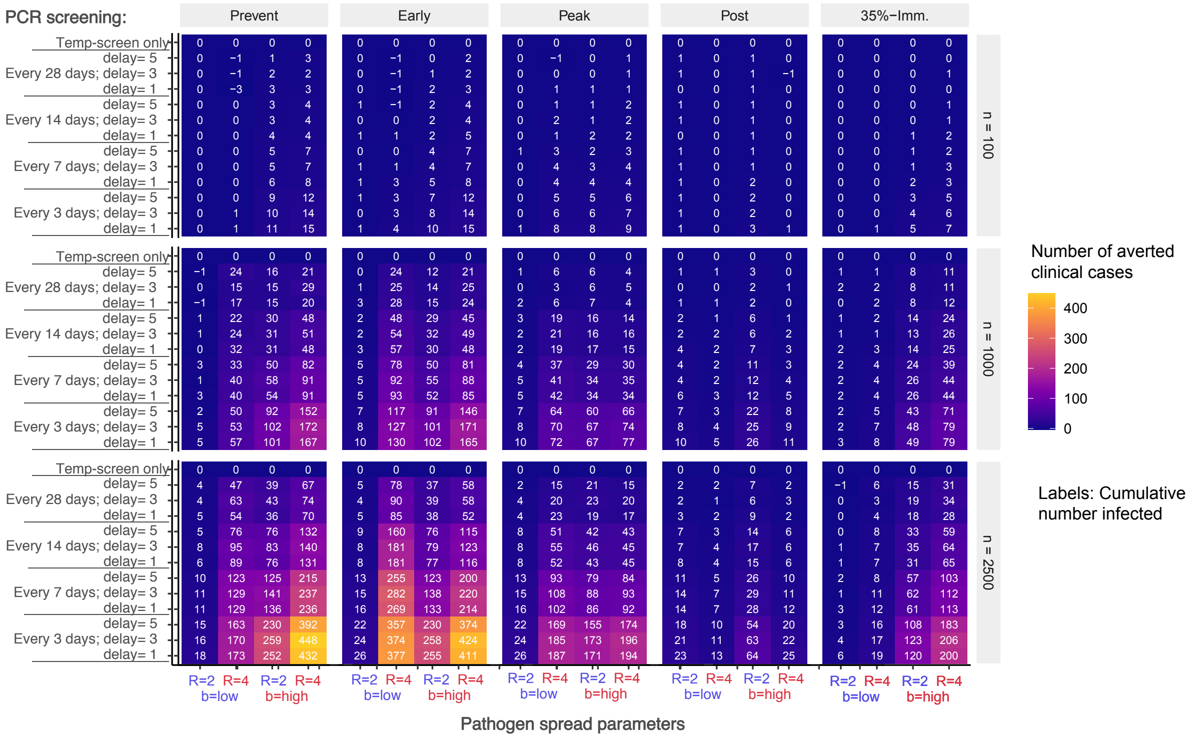

**Supplementary Figure S12.** Proportion of expected clinical cases averted by PCR-screening relative to baseline of temperature screening alone when only 75% of workers participate in the screening program. Results are stratified according to stage at which PCR-screening is implemented (Prevention, Early, Peak, Post, 35% Immune), workforce size (n = 100, 1000, 2500), and high/low workforce *R* and background transmission rates. Absolute number of expected clinical cases under the baseline is labeled in the top line of each colored block, and each cell label shows the clinical cases relative to baseline ($Cases_{scenario}$ / $Cases_{baseline}$). Colored shading indicates the cumulative proportion of the workforce that experiences clinical disease.

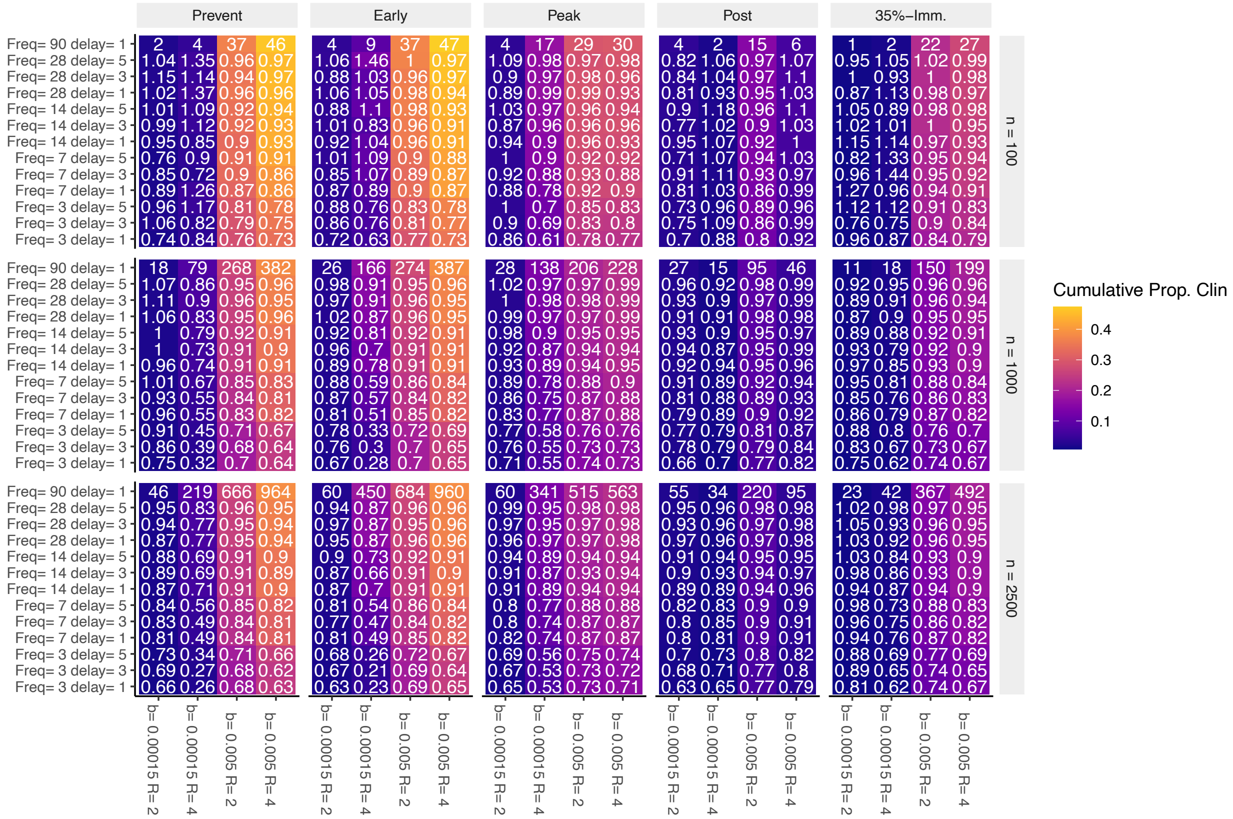

**Supplementary Figure S13.** Cumulative number of additional infections when only 75% of workers participate in the screening program. Results are stratified according to stage at which PCR-screening is implemented (Prevention, Early, Peak, Post, 35% Immune), workforce size (n = 100, 1000, 2500), and high/low workforce *R* and background transmission rates. Each cell label shows the absolute number of additional infections. Colored shading indicates the cumulative proportion of the workforce that is infected.

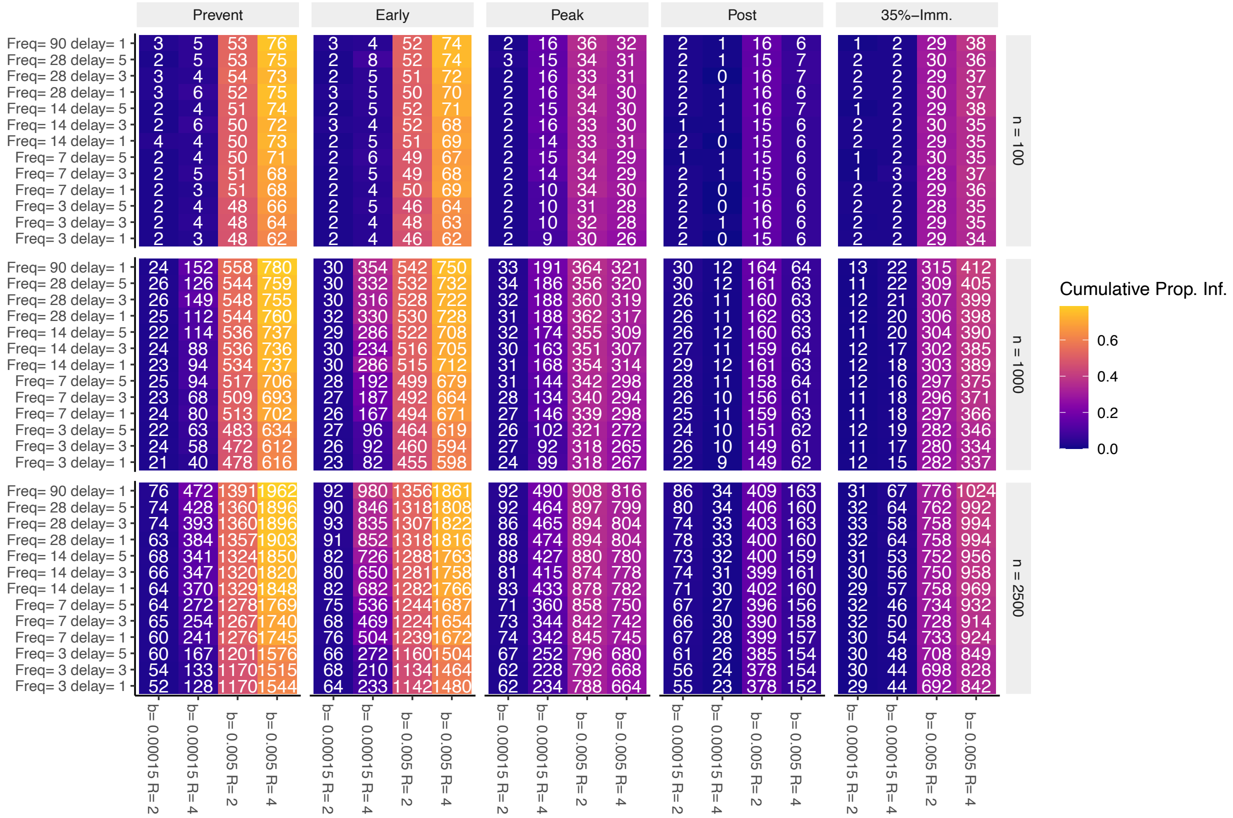

**Supplementary Figure S14.** Cumulative number of additional clinical cases when only 75% of workers participate in the screening program. Results are stratified according to stage at which PCR-screening is implemented (Prevention, Early, Peak, Post, 35% Immune), workforce size (n = 100, 1000, 2500), and high/low workforce *R* and background transmission rates. Each cell label shows the absolute number of additional clinical cases. Colored shading indicates the cumulative proportion of the workforce that experiences clinical disease.

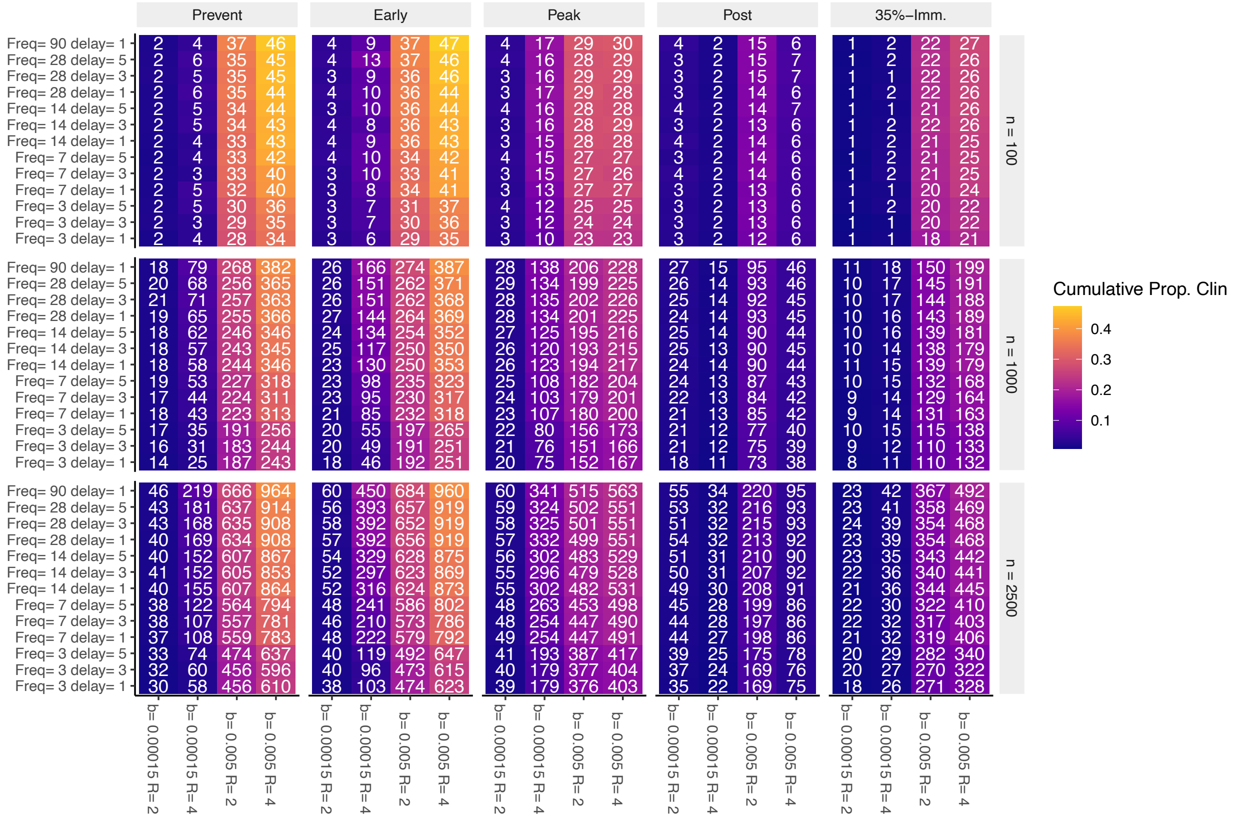

**Supplementary Figure S15.** Proportion of infections due to workplace transmission when only 75% of workers participate in the screening program. Results are stratified according to stage at which PCR-screening is implemented (Prevention, Early, Peak, Post, 35% Immune), workforce size (n = 100, 1000, 2500), and high/low workforce *R* and background transmission rates. Each cell label shows the cumulative number of additional workplace transmission events. Colored shading indicates the proportion of all transmission events that were attributed to the workplace.

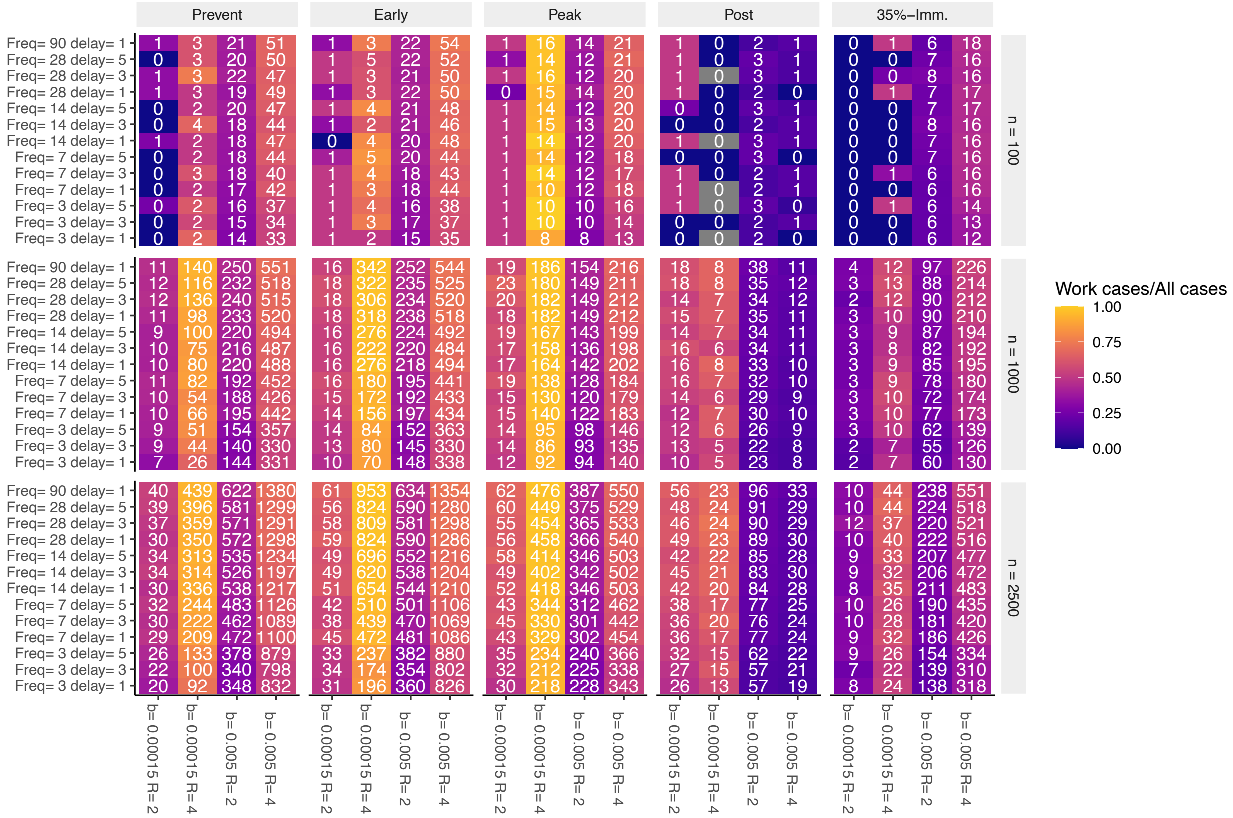

**Supplementary Figure S16.** Number of infections detected via PCR-screening when only 75% of workers participate in the screening program. Results are stratified according to stage at which PCR-screening is implemented (Prevention, Early, Peak, Post, 35% Immune), workforce size (n = 100, 1000, 2500), and high/low workforce *R* and background transmission rates. Each cell label shows the absolute number of infections detected via PCR-screening. Colored shading indicates the proportion of infections that were detected.

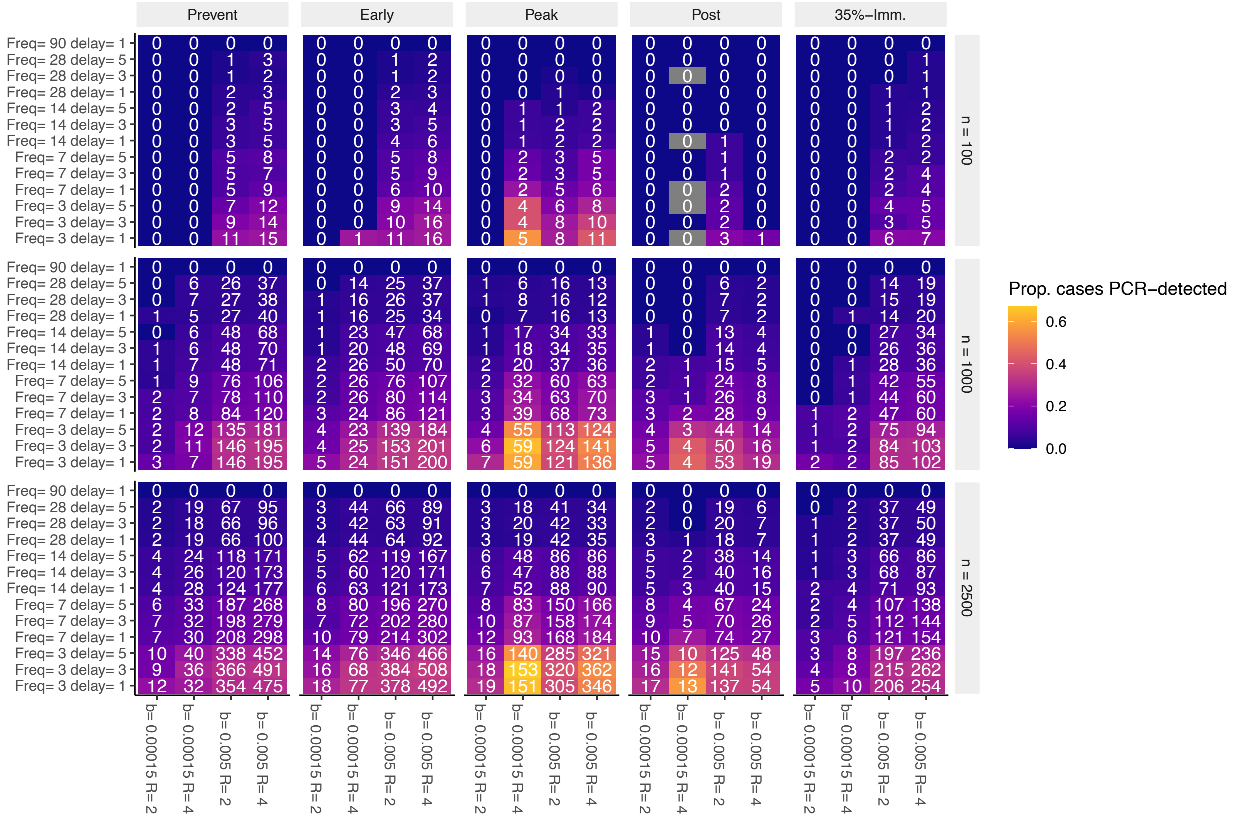
